# Immunogenomic landscape of T cell repertoire after human lung transplantation and its clinical significance

**DOI:** 10.1101/2025.06.03.25328904

**Authors:** Wenyu Jiao, Katherine D. Long, Tyla Young, Constanza Bay Muntnich, Adriana Prada Rey, Morcel Khwajazadah, Joshua H. Wang, Vineetha Mohan, Kortney Rogers, Arnold Valena, Joseph Costa, Luke Benvenuto, Joshua Sonett, Philippe Lemaitre, Frank D’Ovidio, Selim Arcasoy, Jianing Fu

## Abstract

Despite advances in surgical techniques and immunosuppression, long-term survival after lung transplantation (LuTx) remains suboptimal due to high rates of rejection, infection and graft dysfunction. To address this, we investigated post-LuTx T cell dynamics—tracking repopulation, clonal distribution, alloreactivity, and microbial reactivity—through flow cytometry and TCRβ sequencing of serial bronchoalveolar lavage (BAL) and peripheral blood samples. Pre-transplant mixed lymphocyte reactions coupled with TCRβ-seq identified alloreactive TCRs in both graft-versus-host (GvH) and host-versus-graft (HvG) directions, while pathogen-reactive clones were defined via cross-referencing with public databases. We observed progressive establishment of a recipient-derived tissue-resident memory T cell (TRM) repertoire in the BAL, stabilized predominantly by pre-existing, multi-tissue-shared TCRs. Clonal BAL–blood overlap was significantly driven by CD8⁺ non-alloreactive recipient TCRs originating from multiple tissues. A higher HvG:GvH TCR ratio correlated with faster recipient T cell repopulation in BAL, and HvG enrichment in BAL (but not peripheral blood) was associated with early rejection and reduced pulmonary function. Pathogen-reactive TCRs expanded in BAL during infection and were enriched within the non-alloreactive repertoire. This comprehensive TCR landscape analysis highlights the dual roles of T cells in maintaining mucosal homeostasis and contributing to rejection or infection pathogenesis. These findings support the development of precise, mechanism-informed diagnostics to better tailor immunosuppression and ultimately improve LuTx outcomes. Additionally, our work establishes LuTx as a powerful model for studying human tissue-adapted immunity, offering novel insights into the establishment, maintenance, and functional specialization of TRM repertoires.

## Introduction

Lung transplantation (LuTx) is the final therapeutic option for patients with end-stage respiratory diseases, including chronic obstructive pulmonary disease, idiopathic pulmonary fibrosis and pulmonary hypertension [1]. Approximately 2,500 to 3,000 LuTx procedures are performed annually in the United States in recent years, predominantly in adults (over 95%) [2]. While short-term outcomes have improved in recent years, long-term survival remains poor, with 5- year survival rate of 60% [2]. Rejection remains the primary complication limiting transplant success, even with lifelong immunosuppression [3–5]. Although most early rejection episodes can be managed by intensifying immunosuppression, these treatments increase infection risk and predispose patients to high-grade late rejections [6], both of which are key risk factors for chronic lung allograft dysfunction (CLAD), the leading cause of mortality post-LuTx [7].

A major challenge complicating clinical management after LuTx is the lack of accurate diagnostic tools for rejection. Acute cellular rejection (ACR), mediated primarily by donor-reactive recipient T cells, is histologically diagnosed by perivascular and interstitial mononuclear infiltrates in lung tissue, graded by inflammation severity in vascular (A0 to A4) and airway (B0 to B2R) compartments [8]. However, histology-based ACR diagnosis suffers from low sensitivity/ specificity, difficulties distinguishing it from infection-driven inflammation, and high interobserver variability [8–10]. Current efforts to identify novel rejection biomarkers [11–14] face a critical challenge: the inability to distinguish pathogenic donor-reactive recipient T cells from protective T-cell subsets, such as microbial-reactive recipient T cells or recipient-reactive donor T cells that combat infection or rejection, respectively. Therefore, distinguishing donor vs. recipient T cells and integrating their functional phenotypes (alloreactive vs. microbial-reactive) with clonotype tracking via TCRβ sequencing offers a promising new approach. This strategy can map T-cell origin, distribution, and participation in alloresponses and mucosal homeostasis post-LuTx, enabling mechanistic insights into rejection and precision diagnostics to optimize immunosuppression.

Our group did extensive studies in the field of T cell immunogenomics of human alloresponses to reveal mechanisms of rejection and tolerance [15–23]. Our data in human intestinal transplantation showed that graft-versus-host (GvH) T cell clones expand in intestinal allografts and migrate into the recipient circulation and bone marrow. These GvH-reactive T cells express high levels of cytotoxic effector genes and mediate lymphohematopoietic GvH responses that attenuate host-versus-graft (HvG) T cell reactions and make space for donor hematopoietic stem and progenitor cell engraftment, promoting mixed chimerism and contributing to possible immune tolerance [15, 17, 18]. Our immune repertoire profiling of T cell clones revealed their dynamic establishment in intestinal allografts and replenishment from the circulating pool in acquiring tissue residency [23]. Like intestines, lung grafts carry large numbers of T cells and antigen-presenting cells (APCs), which may lead to bidirectional (GvH and HvG) alloresponses, the balance of which is expected to determine graft outcomes. Previous LuTx studies frequently do not distinguish donor cells from recipient cells, leading to controversies and difficulties in interpreting their clinical significance [11–13]. Even in studies demonstrated a dynamic replacement of donor T cells by the recipient T cells in lung allografts, followed by the gradual acquisition of a tissue resident memory (TRM) phenotype by graft-infiltrating recipient T cells [24, 25], critical features of donor and recipient T cells are still largely unexplored, including their clonal distribution, alloreactivity and microbial-reactivity.

To determine how functional TCR clones influence clinical outcomes after LuTx, we conducted longitudinal profiling of donor- and recipient-derived T cells in bronchoalveolar lavage (BAL), transbronchial biopsies (TBBx), and peripheral blood samples. This approach enabled comprehensive characterization of the evolving immunogenomic landscape of T cell repertoires following LuTx at multiple levels: (1) chimeric composition, (2) phenotypic evolution, (3) pre-transplant (pre-Tx) tissue origin, and (4) alloreactive and microbial-reactive clonal dynamics.

BAL-to-blood comparisons revealed how tissue-resident clones emerge over time and are associated with rejection, infection, and pulmonary dysfunction. Our study also offers a unique window into the establishment of the human TRM repertoire, uncovering evolutionary hallmarks—such as preferential persistence, phenotypic convergence, and functional compartmentalization—that inform broader principles of tissue-adapted immunity beyond the transplant context.

## Result

### Multilineage chimerism in blood and BAL post-Tx

A total of 13 adult LuTx recipients have been enrolled and transplanted in our cohort since November 2020 (Table S1). COVID-19 affected sample collection for several earlier enrolled patients (Pts1–6). Pts 2 and 5 were excluded from the analysis due to early death on post- operative day (POD)166 and POD16, respectively. Subsequent analysis included 11 patients: 5 female and 6 male patients aged 28 to 70 (median: 63) years at the time of transplant, who received either single LuTx, double LuTx, or liver Tx + double LuTx (Table S1). We performed flow cytometry to distinguish donor- and recipient-derived cells from T/B cell/monocyte lineages (Fig. S1A) in post-Tx peripheral blood mononuclear cell (PBMC) and BAL samples using HLA-allotype specific antibodies (Table S2) as described previously [26]. BAL is commonly used as a surrogate to monitor the immune changes in the lung allograft [24, 25]. Recipient CD14^+^ monocytes exhibited rapid infiltration into BAL compared to recipient CD3^+^ T cells (Fig. S1B), suggesting their potential role as APCs in priming GvH-reactive donor T cell expansion in BAL early post-Tx, consistent with our data in human intestinal transplantation [15]. A small fraction of donor T cells carried by lung graft mucosa or lymphoid tissues migrated into circulation, resulting in low levels of donor T cell chimerism in blood (<3%) (Fig. S1C). Recipient T cell reconstitution kinetics in BAL reached 50% repopulation by POD150, with notable interpatient heterogeneity (Fig. S1C). These dynamics were comprehensively evaluated for associations with bidirectional alloresponses and graft outcomes in the final Results section. B cell findings will be reported in a separate manuscript and are therefore not discussed here.

### Graft-infiltrating recipient T cells gradually establish a stable repertoire with TRM phenotypes in the lung allograft, a stability primarily driven by pre-existing recipient TCRs

In line with previous report [24], we evaluated the TRM features of CD8 T cells in post-Tx BAL by flow cytometry and found that donor cells overall maintained as TRMs (Fig. 1A, 1B), while graft-infiltrating recipient cells gradually obtained TRM features by upregulating expression of CD69 and CD103 (Fig. 1A, 1B). To investigate the relationship between tissue residency and T cell clonal stability, we calculated the Jensen-Shannon Divergence (JSD) values (Fig. 1C) between TCR repertoires from BAL and PBMC samples collected at sequential post-Tx timepoints in each patient with TCRβ-seq data available (Fig. S2). Notably, JSD values in BAL significantly declined over time (r=-0.5285, p=0.0046), whereas those in PBMCs remained stable (r=-0.3035, p=0.1402). This suggests the progressive establishment of a recipient-derived TRM pool with enhanced repertoire similarity and stability in the lung allograft but not in the periphery (Fig. S2). To evaluate the impact of sampling intervals, we analyzed JSD values against ΔPOD (time between adjacent samples) and found no association with longer intervals, indicating that timing variability did not significantly bias our interpretation (Fig. S3).

**Fig. 1.**
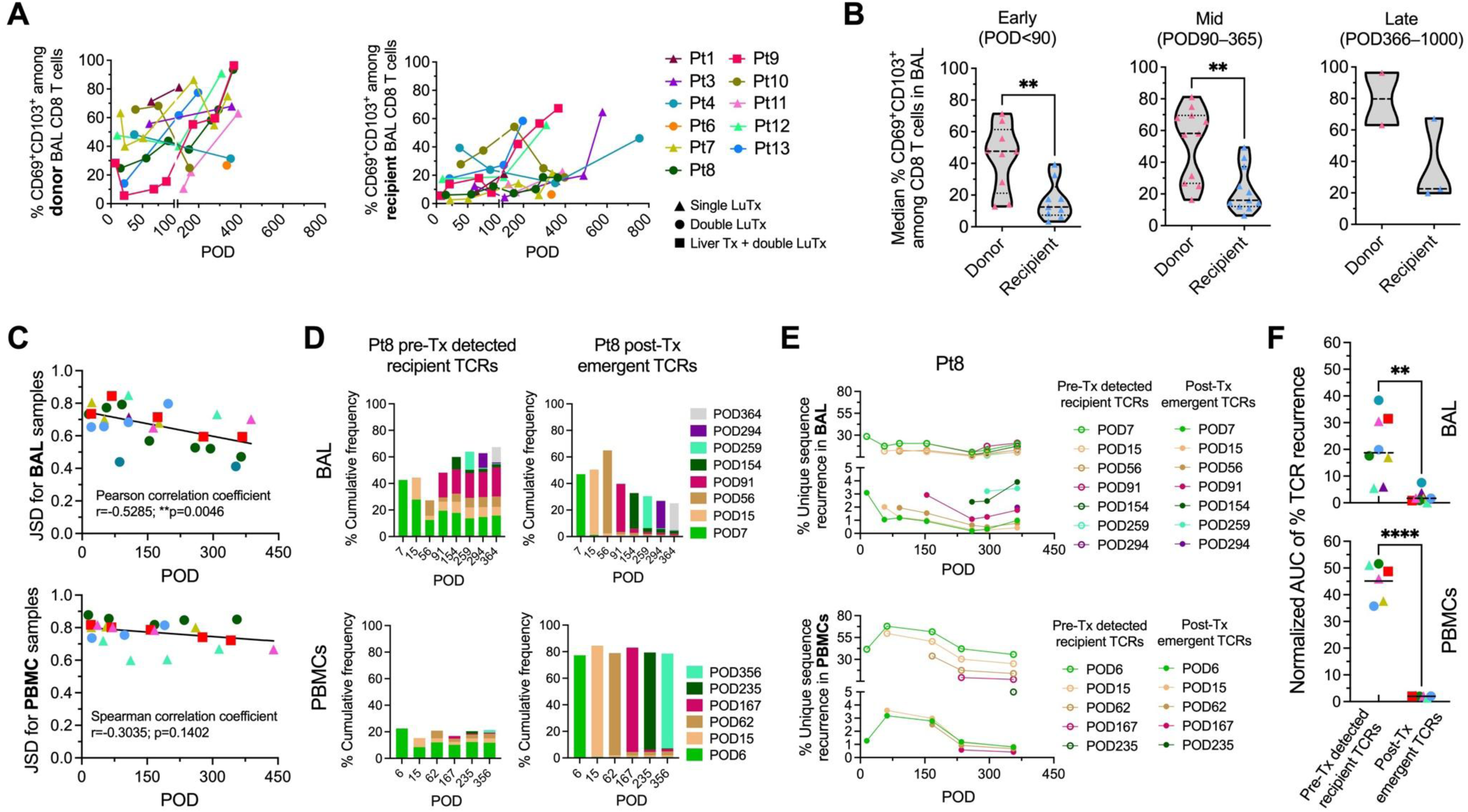
Dynamic establishment of recipient TCR repertoires in BAL and PBMCs. (A) Percentages of donor and recipient CD8+ T cells with TRM phenotypes (CD69+ CD103+) among all the patients (n=11) who underwent post-Tx BAL sample analysis by flow cytometry. (B) Comparison of donor and recipient CD8+ T cell percentages with TRM phenotypes. Samples were stratified into three groups based on postoperative day (POD). (C) Jensen-Shannon Divergence (JSD) values between adjacent sampling time points in BAL and PBMC from patients’ (n=8). (D) Pre-Tx-detected recipient TCRs and post-Tx newly detected TCRs contributed to the TCR repertoires of BAL and PBMCs in Pt8. All the per-Tx samples (listed in Fig. S2), including BAL, lung, PBMC, lung-associated lymph nodes (LLN), and spleen, were used to define the pre-Tx-detected TCRs. In panel D and Fig. S4, pre-Tx-detected donor TCRs (data not shown), pre-Tx-detected recipient TCRs, and post-Tx newly detected TCRs collectively account for 100% of each sample. (E) Recurrence proportions of TCR subsets (defined in D) by unique sequence count (unweighted) post-Tx. Each TCR subset (except those detected at the last sampling time) has a corresponding recurrence proportion group. (F) Comparison of recurrence between the earliest pre-Tx-detected recipient TCR subset and the earliest post-Tx newly detected TCR subset per patient. Recurrence values were normalized by the area under curve (AUC) divided by the time span (shown in E and Fig.S4C). Unpaired t test was performed for panel B. Wilcoxon test and Paired t test were performed for the upper and lower plots, respectively. Solid horizontal bars indicate means, and dotted horizontal bars indicate medians. **p < 0.01, ****p < 0.0001.

To delineate mechanisms underlying post-Tx BAL repertoire stabilization, we partitioned TCRs detected in post-Tx samples into pre-existing (pre-Tx detected) donor and recipient TCRs, and "*de novo*" (post-Tx emergent) clonotypes. These were subclassified by initial detection timepoint and assessed for clonal persistence via recurrence rates of unique CDR3 sequences (Figs. 1D, S4). Pre-existing donor TCRs were not discussed further in this analysis given overall less dominant presence in post-Tx samples, although demonstrate considerable levels of clonal recurrence in post-Tx BAL (data not shown), in line with their TRM phenotypes. We observed that pre-Tx recipient TCRs detected across multiple PODs progressively accumulated in both BAL and PBMC repertoires, with many TCR sequences identified at early point(s) recurring at later stages (PBMCs: mean 30.32% ± 18.90%; BAL: mean 17.55% ± 10.04%). In striking contrast, while newly detected post-Tx TCRs constituted a substantial proportion of both the BAL and PBMC repertoires, only a minor fraction demonstrated recurrent detection (PBMCs: mean 1.27% ± 1.38%; BAL: mean 1.77% ± 2.30%) (Figs. 1E, S4). Quantitative analysis using normalized area under curve (AUC) metrics showed pre-Tx recipient TCRs (first detected at the earliest POD) had significantly higher recurrence rates than post-Tx newly detected TCRs in both BAL and PBMC compartments (Fig. 1F). This demonstrates that repertoire stability post-Tx was primarily maintained by the pre-existing recipient TCRs rather than newly detected clones. Among pre-Tx recipient-derived TCRs, we observed that CD8 TCRs consistently demonstrated higher recurrence rates than CD4 TCRs in both BAL and PBMC compartments (Fig. S5).

### Cross-tissue pre-existing recipient TCR sequences exhibited enhanced clonal recurrence in both BAL and blood compartments post-Tx

To delineate how pre-Tx recipient tissue origins (Table S3) shape post-Tx repertoire establishment, we systematically categorized recipient TCRs detected in post-Tx BAL and PBMC samples into seven groups based on their pre-Tx detection patterns: (1) lymphoid- PBMC-lung shared, (2) PBMC-lung shared, (3) lymphoid-lung shared, (4) lymphoid-PBMC shared, (5) lung only, (6) PBMC only, and (7) lymphoid only. Notably, while single-tissue-derived TCRs (groups 5–7) constituted substantial fractions of pre-Tx repertoires, their representation was markedly reduced in both post-Tx BAL and PBMCs (Figs. 2A, S6). Conversely, multi-tissue shared TCRs (groups 1–4) – particularly triple-shared clones (group 1) – dominated the post-Tx repertoires (Figs. 2A, S6).

**Fig. 2.**
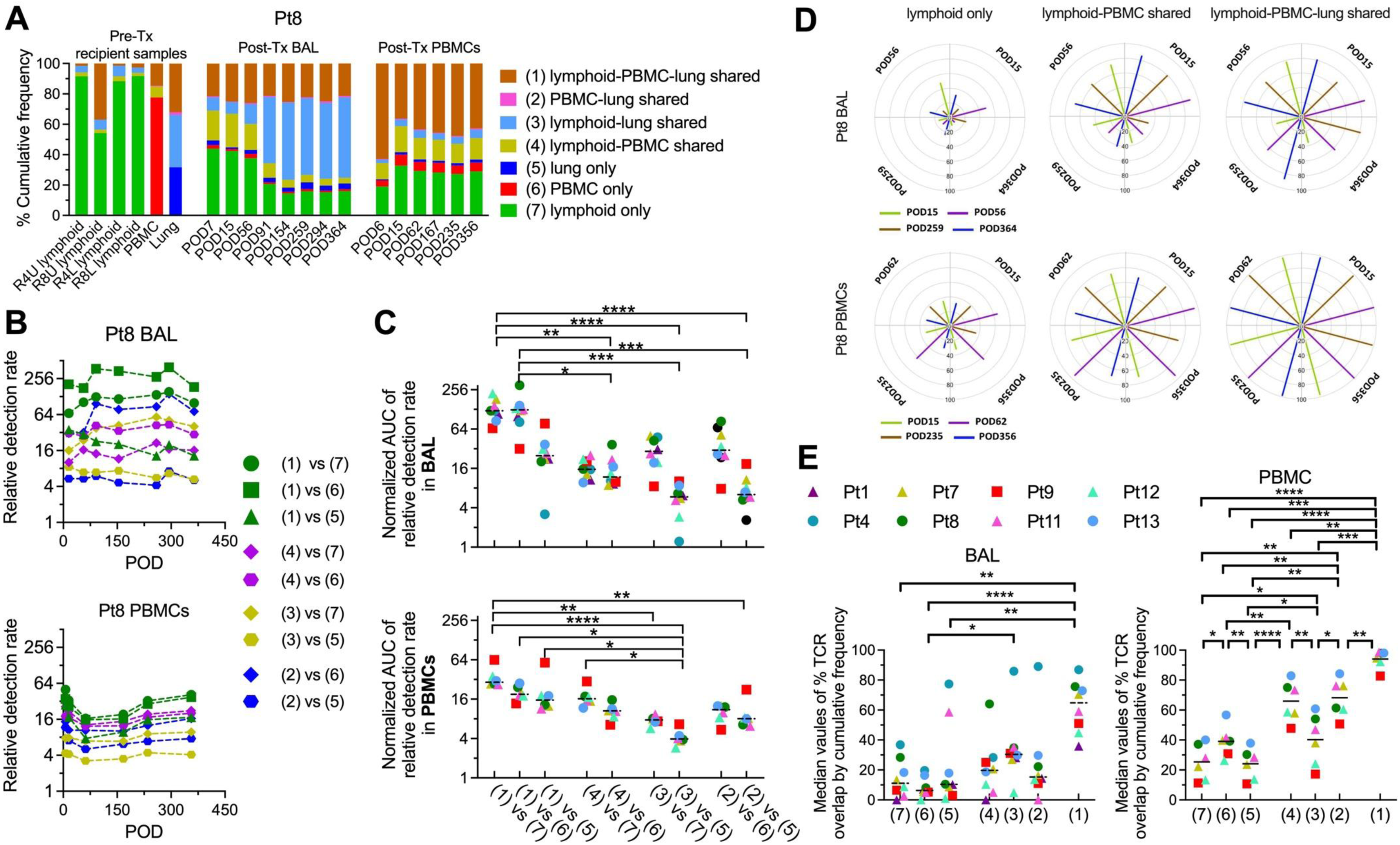
Composition, relative detection rate, and persistence of T cell subsets by tissue origin. (A) Cumulative frequencies of recipient TCRs classified as “lymphoid-PBMC-lung shared” (1), “PBMC-lung shared” (2), “lymphoid-lung shared” (3), “lymphoid-PBMC shared” (4), “lung only” (5), “PBMC only” (6), and “lymphoid only” (7) in pre-Tx and post-Tx samples. R4U: recipient CD4 unstimulated; R8U: recipient CD8 unstimulated; R4L: recipient CD4 CFSE ^low^; R8L: recipient CD8 CFSE^low^. (B) Relative detection rates in post-Tx Pt8 BAL and Pt8 PBMC for sequences identified pre-Tx. The relative detection rate (odds ratio) compares the detection rate of two sequence subsets. A value > 1 indicates a higher detection of the first subset versus the second. (C) Time-normalized AUC of all relative detection rate values. (D) Representative circle plots of TCR subsets in Pt8 BAL and PBMC: “lymphoid only” (non-shared TCRs), “lymphoid- PBMC shared” (double shared TCRs), and “lymphoid-PBMC-lung shared”. Colored line lengths represent the % overlap (cumulative frequency, 0–100) between the sampling timepoint (line) and the reference timepoint (outside circle label) The latter serves as the denominator. (E) Median overlap % among TCR subsets, stratified by pre-Tx detection, across all patients’ post- Tx BAL and PBMC samples. The Friedman test followed by Dunn’s multiple comparisons test was performed for panels C and E. Solid horizontal bars indicate means and dotted horizontal bars indicate medians. *p < 0.05, **p < 0.01, ***p < 0.001, ****p < 0.0001.

We quantified the relative detection rates of multi-tissue shared TCRs vs. non-shared TCRs in post-Tx BAL and PBMC samples. In our representative patient analysis, shared TCR subsets, particularly lymphoid-PBMC-lung triple-shared TCRs, demonstrated significantly higher detection rates compared to non-shared TCRs (Fig. 2B). This pattern was consistent across all patients when assessed via normalized AUC analysis of detection rate kinetics in both BAL and PBMC samples, with triple-shared TCRs exhibiting the highest relative detection rates in the BAL compartment (Fig. 2C).

To quantify the temporal similarity of TCR repertoires across tissue origins, we assessed sequence overlap (templated-weighted) among non-shared, double-shared, and triple-shared TCR subsets of post-Tx BAL and PBMC samples (representative patient: Fig. 2D). Pooled analysis across all patients revealed significantly higher median overlap proportions in triple-shared vs. double-shared or non-shared subsets (Fig. 2E). While PBMC repertoires exhibited a strict hierarchical overlap pattern (triple- > double- > non-shared), BAL samples showed a similar but less pronounced trend. Collectively, our results demonstrate that TCRs detected in multiple pre-Tx recipient tissues - including peripheral blood, lung-associated lymphoid tissue, and lung parenchyma - exhibit superior engraftment potential in both BAL and blood compartments. These multi-tissue-derived TCRs showed significantly higher recurrence rates and were the primary drivers of post-Tx TCR repertoire stability.

### T cell exchange between blood and BAL persists over one-year post-Tx, primarily mediated by both multi-tissue-derived recipient TCRs present prior to transplantation and recipient non-alloreactive CD8 TCRs

To elucidate the spatial dynamics of TCR repertoire distribution post-LuTx, we systematically analyzed T cell crosstalk between PBMC and BAL compartments (Fig. 3A) across three post- operative periods: early (POD<90), mid (POD90-365), and late (POD>365). Using a comparison matrix (Fig. 3B), we quantified the proportion of BAL TCR sequences detectable in corresponding PBMC samples. While no significant temporal pattern emerged in overlap fractions across periods (Fig. 3B), a persistent T cell crosstalk between peripheral blood and lung allografts was observed throughout the first year, with occasionally lower early-phase overlap (e.g., groups A, B and C in Pt9) potentially attributable to residual donor T cells in grafts. Aggregate analysis (compiled data points from groups A to I for each patient) revealed that BAL-PBMC clonal overlap was significantly enriched for recipient-derived pre-Tx TCRs (Fig. 3C), particularly triple-shared (lymphoid-PBMC-lung) TCRs (Fig. 3D) and CD8 non-alloreactive subsets (CD8 nonHvG) (Fig. 3E). These findings implicate tissue-experienced, non-alloreactive CD8 T cells act as the primary migratory population and key mediators of systemic-lung immune surveillance post-LuTx.

**Fig. 3.**
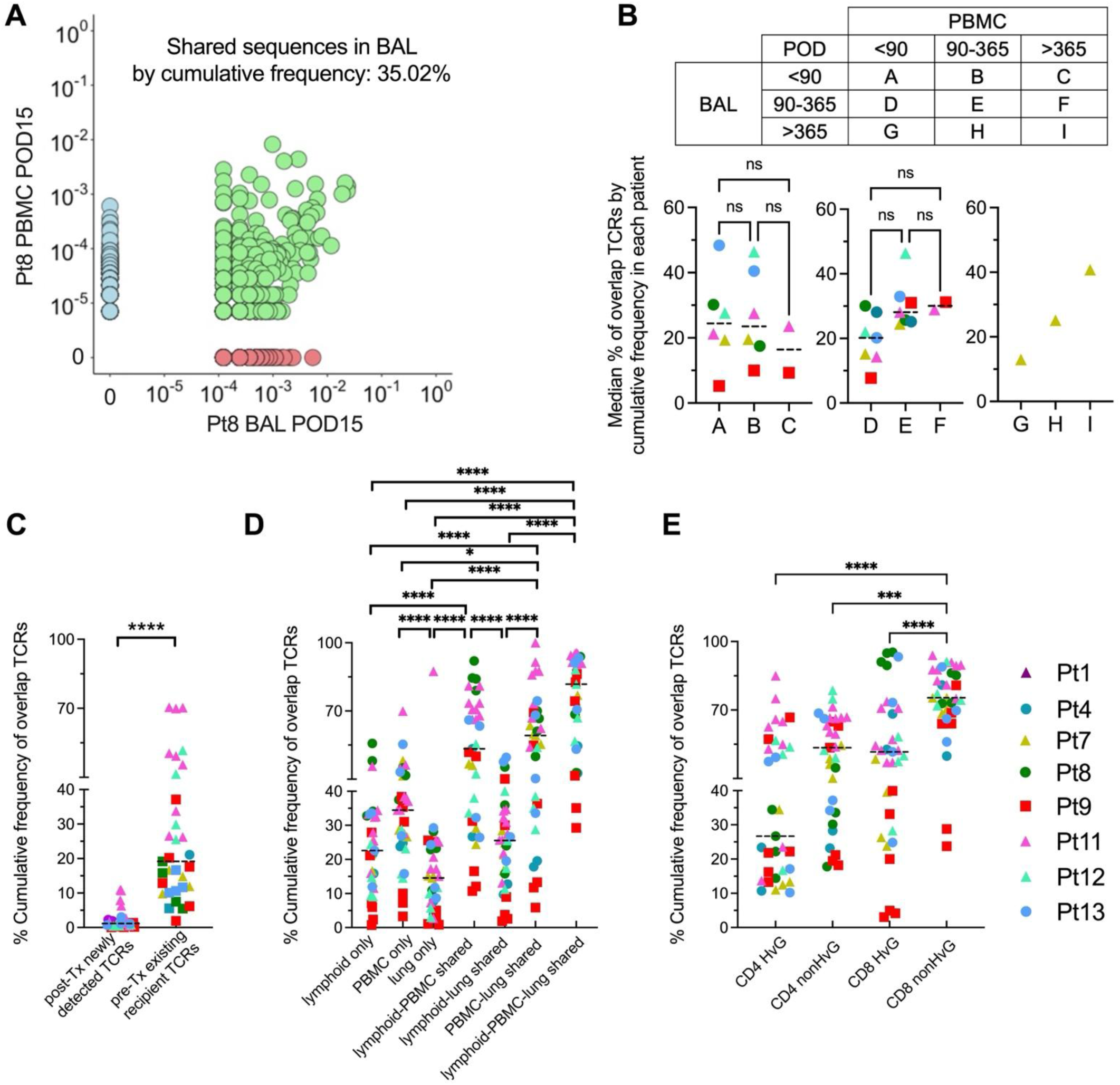
Crosstalk between PBMC and BAL samples and the differences among TCR subsets. (A) Representative correlation plots of clone frequencies between paired BAL and PBMC samples from Pt8. (B) BAL and PBMC samples were grouped into 9 pairs based on sampling dates. The median cumulative frequency of shared sequences in BAL of each patient in each group were summarized and compared across early (group A, B, C), mid (group D, E, F) and late (G, H, I) periods of BAL sampling. (C) Comparison of PBMC-BAL overlapping percentages between pre-Tx-detected recipient TCRs and post-Tx newly detected TCRs when compiled all 9 pairs (defined in panel B). (D) Comparison of PBMC-BAL-overlapping percentages among pre-Tx detectable recipient TCR subsets by tissue origin (defined in Fig. 2) when compiled all 9 pairs (defined in panel B). (E) Comparison of PBMC-BAL overlapping percentages among CD4 and CD8 HvG and nonHvG TCRs when compiled all 9 pairs (defined in panel B). Kruskal-Wallis test followed by Dunn’s multiple comparisons test was performed for panel B. Wilcoxon test was performed for panel C. Friedman test followed by Dunn’s multiple comparisons test were performed for panels D and E. Dotted horizontal bars indicate medians. *p < 0.05, ***p < 0.001, ****p < 0.0001.

### Microbial-reactive TCRs were enriched in lung allografts during infection and predominantly composed of non-alloreactive T cell populations

The requirement for lifelong immunosuppression in LuTx recipients significantly increases infection susceptibility. In our cohort, all patients experienced ≥1 pathogen infection, with many developing recurrent infections by diverse pathogens (Fig. 4A, Table S1). We hypothesized that non-alloreactive T cells (defined by non-reactivity to donor or recipient antigens in pre-Tx allogeneic MLRs) may persist and expand pathogen-specific clones during infection. Using a strategy we previously validated in human intestinal transplantation [23], we systematically identified microbial-reactive TCRs by cross-referencing our dataset with published TCR databases (including COVID-19-specific clones) [27–32], focusing on CDR3β (V-J-AA) signatures. In patients with post-Tx EBV or CMV infection - the most clinically significant viral pathogens - we observed significant enrichment of EBV- or CMV-reactive TCRs in BAL and lung allografts during active infection vs. peripheral blood (Figs. 4A, 4B). These findings demonstrate that our TCR mapping approach reliably tracks pathogen-specific responses. To comprehensively characterize microbial-reactive TCRs and assess their potential alloreactive cross-reactivity, we performed detailed annotation of TCR specificity in Pt13 (as a representative case), categorizing TCR clones as alloreactive (CD4/CD8 HvG, CD4/CD8 GvH) and non-alloreactive (CD4/CD8 nonHvG, CD4/CD8 nonGvH). This analysis encompassed TCRs reactive to common bacterial/viral pathogens (Fig. 4C) and COVID-19 (Fig. S7A) in post-Tx lung, BAL, and/or PBMCs. Pooled analysis across our cohort revealed a consistent pattern: both donor- and recipient-derived non-alloreactive T cell clones demonstrated significantly greater enrichment for microbe-reactive TCRs compared to their alloreactive counterparts. This microbial-focused specialization was observed universally across all tissue compartments (lung allograft, BAL and peripheral blood), suggesting non-alloreactive clones primarily mediate pathogen surveillance (Figs. 4D, S7B).

**Fig. 4.**
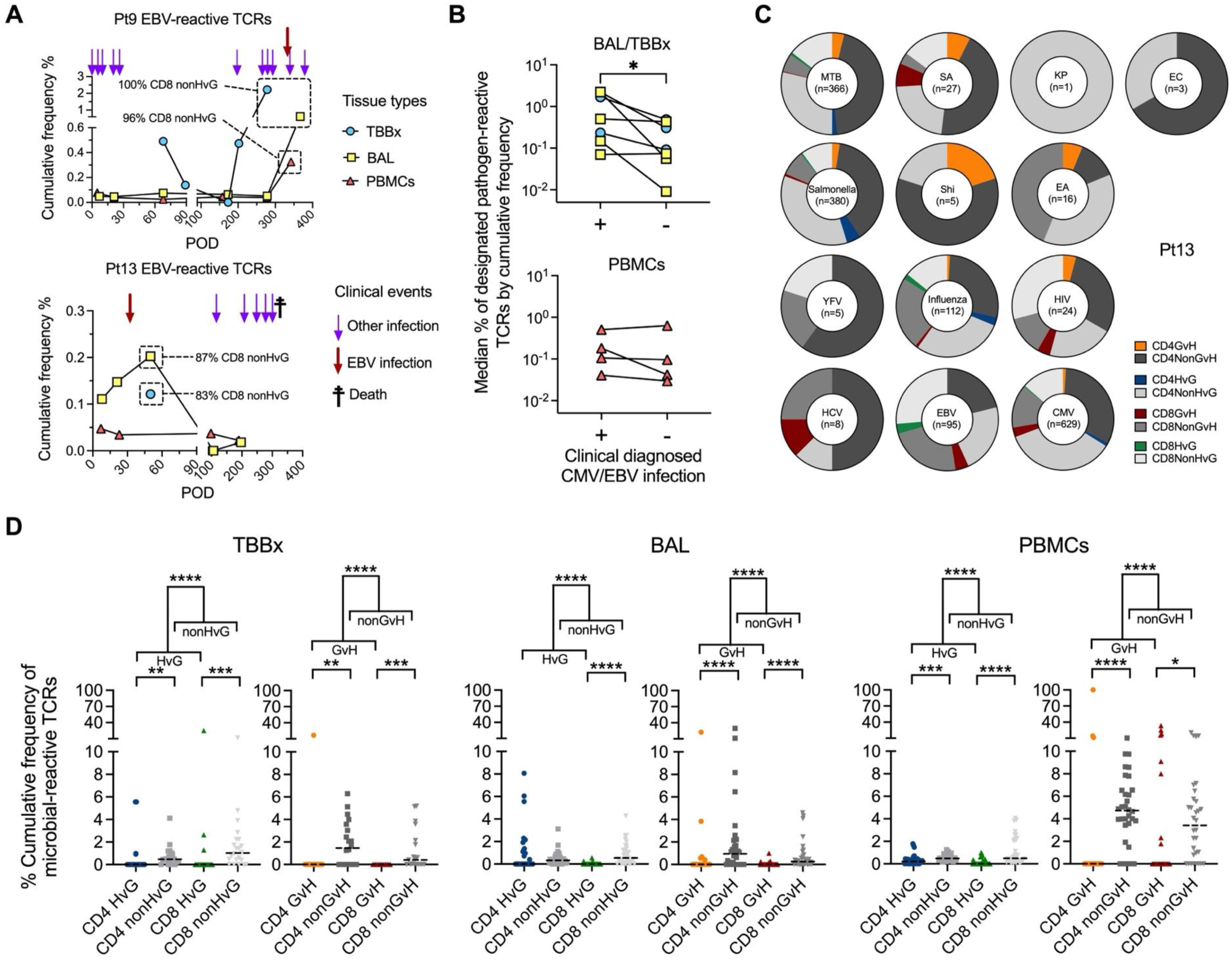
Pathogen-driven TCR proportions and reactivity among TCR subsets with different alloreactivities. (A) Cumulative frequency proportion of EBV-reactive TCRs in post-Tx TBBx, PBMC and BAL samples of Pt9 and Pt13. The proportion of pre-Tx defined CD8 nonHvG TCRs among EBV-reactive TCRs in designated samples were annotated in the plots. (B) Median cumulative frequency (%) of designated pathogen-reactive TCRs in local (lung/ BAL) versus circulating compartments from 4 applicable patients with or without specific infection (CMV/EBV). "With” infection" samples were collected within ± 30 days of clinical diagnosis. (C) Composition of pathogen-reactive TCRs in Pt13 at the unique sequence level subgrouped by alloreactive vs nonalloreactive identification in pre-Tx MLRs, including CD4 and CD8 GvH and nonGvH, HvG and nonHvG categories. Pie chart centers indicate total unique TCRs reactive to each pathogen in all samples sequenced from Pt13. (D) Cumulative frequency of pathogen- reactive TCRs in post-Tx samples (n=8 patients), stratified by alloreactive (CD4 HvG, CD8 HvG, CD4 GvH, CD8 GvH) vs nonalloractive (CD4 nonHvG, CD8 nonHvG, CD4 nonGvH, CD8 nonGvH) identification. Wilcoxon tests were performed for panels B and D. Dotted horizontal bars indicate medians. *p < 0.05, **p < 0.01, ***p < 0.001, ****p < 0.0001. EBV: Epstein-Barr virus; CMV: cytomegalovirus; MTB: mycobacterium tuberculosis; SA: staphylococcus aureus; KP: klebsiella pneumoniae; EC: Escherichia coli; Shi: shigella; EA: Enterobacter aerogenes; YFV: yellow fever virus; HIV: human immunodeficiency virus; HCV: hepatitis C virus.

### HvG-GvH imbalance drives recipient T cell repopulation dynamics in BAL and HvG- reactive TCR enrichment in BAL correlates with early rejection and impaired pulmonary function

In line with our study in human intestinal transplantation [18], we hypothesize that the balance between GvH and HvG alloresponses may determine graft outcomes following LuTx. GvH- and HvG-reactive TCR clones were defined using pre-Tx MLRs and high-throughput TCRβ sequencing, as we previously described [15, 18]. Pre-Tx MLRs utilized donor and recipient cells isolated from lung-associated lymph nodes (LLNs). We monitored GvH- and HvG-reactive TCRs in post-Tx BAL samples from 8 patients with available pre-Tx specimens (Figs. 5A, S8A, S5).

**Fig. 5.**
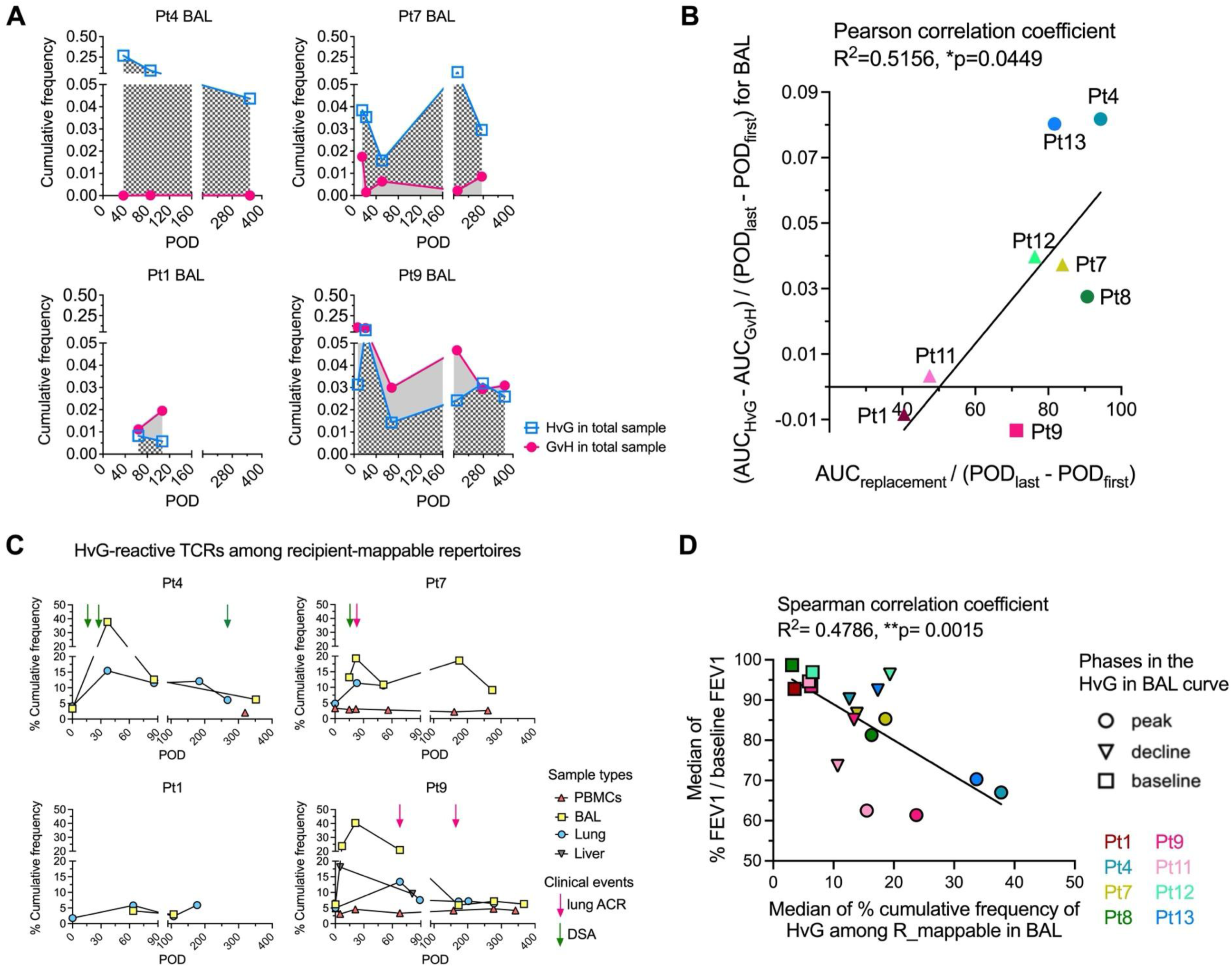
Relationship between TCR repertoire features and clinical outcomes. (A) Cumulative frequency of GvH and HvG clones in BAL specimens from representative patients. (B) Normalized area under the curve (AUC) analysis of GvH and HvG clones across all patients (Y- axis), with normalization by days of measurement (POD_last_ - POD_first_). The X-axis shows similarly normalized AUC values for recipient T cell replacement (from Fig. S1C). (C) Percentage cumulative frequency of HvG-reactive sequences among recipient mappable repertoires in post- Tx PBMC, BAL and lung or liver allografts in representative patients. (D) According to the HvG in BAL over POD curves (Figs. 5C, S8B), three phases were defined as peak (circle), decline (reverse triangle) and baseline (square) (Fig. S8C) and the median of % cumulative frequency of HvG among recipient mappable repertoires in BAL was calculated for each patient (x axis). This is further associated with the pulmonary functions reflected by the median of % FEV1/baseline FEV1 values of each patient (y axis). FEV1: forced expiratory volume in one second. Pearson correlation analysis was performed for panel B (data follow normal distribution). Spearman correlation analysis was performed for panel D (data did not follow normal distribution). *p < 0.05, **p < 0.01.

Five patients (Pts 4/7/8/12/13) with faster recipient T cell repopulation in the BAL (Fig. S1C) showed dominant HvG- over GvH-reactive clones, which is different than the remaining 3 patients (Pts1/9/11) with slower recipient T cell repopulation in BAL (Figs. 5A, S8A). A statistically significant correlation was identified between rapid recipient T cell repopulation and a dominant ratio of HvG- to GvH-reactive TCRs in BAL (Fig. 5B), which were quantified by AUC normalized by days of measurement for both the replacement rate and HvG vs. GvH alloreactivities in the BAL. A similar trend was observed in lung transbronchial biopsies (TBBx: data not shown). Instead of looking at total TCR repertoires (Fig. 5A-B), we now focused on recipient mappable TCR repertoires (Figs. 5C-D, S8B, Table S5), defined as TCRs detectable in pre-Tx unstimulated or MLR CFSE^low^ T cell populations from recipient LLNs. These repertoires include both HvG and nonHvG TCRs. During early ACR in lung allografts (Pt7 POD22, Pt9 POD68), we observed high levels of HvG TCRs among recipient mappable repertoires in BAL and, to a lesser extent, TBBx, but not in PBMCs (Fig. 5C), indicating local HvG clonal expansion correlates with early rejection. In liver allografts of Pt9, higher HvG TCR levels were seen during mild/moderate ACR (POD5) vs. minimal ACR (POD81) (Fig. 5C). In some patients, early post-Tx (within POD90) HvG peaks were identified in BAL, but not PBMCs, despite negative histopathology for ACR, but preceded late complications (e.g., late ACR [POD>200 in Pts 8, 13], *de novo* DSA [Pts 4,11,13], CLAD [Pt4], or death [Pt13 POD303, Pt4 POD1556]), suggesting undiagnosed subclinical rejection and the need for immunosuppression refinement (Figs. 5C, S8B).

To investigate the relationship between HvG responses and pulmonary function despite temporal mismatches in measurement timing, we stratified the post-Tx course into three distinct phases based on BAL HvG TCR dynamics: peak, decline and baseline (Figs. 5C, S8B, S8C).

We then analyzed corresponding forced expiratory volume in one second (FEV1) measurements (normalized to baseline values) within each phase (Fig. S8C). Strikingly, we observed a significant inverse correlation between HvG TCR levels and pulmonary function across phases. These findings establish graft-infiltrating recipient HvG-reactive T cells as potential mediators of rejection and determinants of allograft function.

## Discussion

LuTx outcomes significantly lag behind those of other solid organ transplants due to at least two interrelated challenges. First, the lack of precise rejection diagnostics forces dangerous clinical trade-offs: delayed treatment leads to sustained inflammation and progressive rejection, while overimmunosuppression heightens infection risk—both accelerating CLAD, the leading cause of post-LuTx mortality. Second, donor-derived lymphocytes within the allograft play a critical role in shaping post-Tx immunity but remain poorly understood. Our study addresses this dual pathophysiology through immunogenomic profiling of both donor and recipient T cells. By temporally and spatially mapping bidirectional alloresponses (HvG and GvH) and microbial immunity, we uncover key mechanisms underlying rejection and infection. These insights directly inform novel strategies to overcome diagnostic limitations and optimize immunosuppression, ultimately bridging the LuTx outcomes gap. Our study also establishes LuTx as a powerful model for human TRM biology, providing unprecedented insight into the establishment, maintenance, and functional specialization of TRM repertoires. By overcoming the critical limitation of longitudinal access to non-lymphoid tissues—a major barrier in human immunology—we reveal conserved principles of tissue-adapted immunity with broad implications beyond transplant science.

We demonstrated the progressive establishment of a recipient-derived TRM repertoire in post-LuTx BAL, with repertoire stability predominantly driven by pre-existing, multi-tissue-shared recipient TCRs. These findings suggest a replenishment model where recipient TCRs migrate from the circulating pool, lymphoid reservoirs, or even lung parenchymal niches. This aligns with emerging evidence of re-circulating TRMs – a paradigm shift in TRM biology supported by both murine and human studies [33–38], where tissue-adapted clones retain limited migratory capacity between mucosal sites and secondary lymphoid organs. Building on our prior work in human intestinal transplantation [23]—where single-cell RNA sequencing revealed distinct transcriptional programs in T cells shared across pre-Tx lymphoid and non-lymphoid tissues vs. tissue-restricted clones—we now establish that these multi-tissue-primed T cells tend to represent a recirculating TRM reservoir. In LuTx, such clones (particularly the lymphoid-PBMC- lung triple-shared subset) dominate graft-resident repertoires and exhibit both tissue adaptability and re-circulating potential to facilitate BAL-blood crosstalk. While recirculating TRMs exist in both CD4+ and CD8+ lineages, their dynamics are context-dependent—varying by tissue site and host environment, as demonstrated in both animal models and human studies [38–43].

Notably, we found that non-alloreactive CD8+ recipient T cells, rather than CD4+, played a more active role in BAL-blood crosstalk and were more enriched for microbial-reactive TCRs. This suggests a predominant recirculating phenotype that contributes to host defense against pathogens. Collectively, these findings refine the ’resident memory’ paradigm by demonstrating that human TRMs occupy a spectrum of tissue fidelity, with multi-tissue-primed non-alloreactive CD8 T cell clones more likely to exhibit strategic circulation to surveil dispersed mucosal sites.

It is important to note that the non-alloreactive TCRs defined in our study are specific to a single donor–recipient pair within our cohort. Indeed, prior work by our colleagues has demonstrated that a very high proportion of human T cells—if not all—possess alloreactive potential against a broad array of allogeneic stimulators, as measured by MLRs [44]. Consequently, it is reasonable to infer that pathogen-reactive T cells may frequently exhibit cross-reactivity to alloantigens presented by diverse MHC combinations, consistent with previous human studies [45]. However, in the context of LuTx, where recipients are typically exposed to a single allogeneic donor (excluding re-transplantation scenarios), we observed that HvG–reactive TCRs—unlike pathogen-reactive TCRs, which were more often classified as nonHvG—were disproportionately enriched in post-Tx BAL and TBBx samples compared to peripheral blood during early rejection. This finding supports the potential for using HvG TCRs in combination with pathogen-reactive TCRs as diagnostic markers to distinguish early rejection from infection-driven inflammation. Such differentiation is frequently missed by histology-based diagnostics, often resulting in inappropriate immunosuppression and life-threatening complications.

Rejection and infection are often interrelated in transplantation. Overimmunosuppression to control rejection increases susceptibility to infection, but the relationship is bidirectional. The concept of ’heterologous immunity’—in which virus-specific T cells can cross-react with alloantigens to enhance alloimmunity—has been documented [45, 46]. Barrier organ transplants, particularly the lungs and intestines, are especially vulnerable due to their extensive and often hidden surface areas, making them prone to infection-related rejection [2, 47]. Beyond barrier organs, studies in rodent skin transplantation [48, 49] and human kidney transplantation [50] have shown that immune responses to commensal organisms and CMV, respectively, can promote graft rejection. These findings underscore the need for effective and balanced control of both infection and rejection to improve clinical outcomes across a wide range of solid organ transplants. The current criteria for diagnosing ACR in lung transplant recipients were originally developed in the 1990s, with the most recent update in 2007 [8, 51]. Given the rapid advancements in next-generation sequencing and the rise of multiomic technologies, the field is primed for the development of more precise, mechanism-driven diagnostic tools. To this end, we have established innovative, multidisciplinary platforms to elucidate the mechanisms of rejection and infection, paving the way for future refinement of diagnostic strategies.

Our study has several limitations. First, the small cohort of LuTx patients precluded meaningful multivariate analyses. Early sample collection was impacted by the COVID-19 pandemic, and as a result, early post-Tx blood samples were not obtained from Pts 1 and 4. In addition, the limited availability of microbial-reactive TCRs in reference databases restricted our ability to map a broader repertoire within our dataset. Nonetheless, our findings establish proof-of-concept that this platform is informative for studying human transplantation immunology. To address the limitations of current microbial-reactive TCR annotation, computational approaches such as TCR similarity algorithms (e.g., GLIPH2) could be leveraged to expand identification, as we previously described [22]. However, functional assays remain the gold standard for confirming pathogen reactivity, which was beyond the scope of this study. The observed association between HvG TCRs and rejection was identified in only a subset of patients, which may be partially explained by the presence of subclinical or histologically undetectable rejection [52]. Importantly, even undiagnosed rejection may contribute to pulmonary function decline. Our findings indicate that lung-infiltrating HvG T cells may adversely affect pulmonary function regardless of clinically overt rejection. While clinical data were not always available or temporally aligned with TCR sequencing, our results suggest that incorporating early pulmonary function tests could enhance the interpretation of TCR-seq data and potentially predict the development of CLAD and mortality (e.g., Pts 4 and 13, Fig. 5D, Table S1). Validation of these findings in an expanded cohort will be necessary to confirm their generalizability and clinical utility. Both our current LuTx data (Fig. 5C: Pt9; Fig. S8B: Pt8) and our previous intestinal transplantation studies [21] found that pre-Tx MLR-defined HvG clones show diminished correlation with late-stage (POD>150) rejection episodes. A promising approach to enhance late rejection monitoring would involve additionally performing MLRs using post-Tx PBMCs or BAL-derived lymphocytes to enable detection of putative *de novo* HvG clones that develop post-Tx, similar to our intestinal transplantation findings [21]. However, such investigation extends beyond the current study scope.

Our data provides immunogenomic insights into the dynamic interplay between T cell alloreactivity and microbial reactivity in LuTx, advancing our understanding of rejection mechanisms and informing the refinement of diagnostic frameworks. By dissecting bidirectional alloresponses and tissue-residency dynamics, this study opens new avenues for future research focused on the development of precision therapies that selectively modulate alloimmune responses while preserving antimicrobial immunity—addressing a critical unmet need in improving LuTx outcomes.

## Methods

### Human subject recruitment and clinical protocols

The study was approved by the Columbia University Institutional Review Board (IRB #AAAS6206 & AAAR2681). All subjects or legal guardians provided their written, informed consent and assent when appropriate. Our cohort enrolled thirteen patients: six single-lung transplant recipients (Pts1, 2, 3, 7, 11, 12), six double-lung transplant recipients (Pts4, 5, 6, 8, 10, 13), and one double-lung-and-liver transplant recipient (Pt9). Because Pt2 and Pt5 passed away less than six months post-Tx and there were limited pre-Tx and post-Tx specimens collected from these patients, they were excluded from cohort analysis. All lung allografts were procured from donors after brain death without the use of Normothermic Regional Perfusion.

In the post-Tx period, rejection was graded following the International Society for Heart & Lung Transplantation (ISHLT) guidelines, with attention to acute rejection and airway inflammation [8]. Acute rejection was graded from none, minimal, mild, moderate, to severe, while small airways inflammation was graded as none, low grade, high grade, or ungradable. ACR is diagnosed by the presence of perivascular and interstitial mononuclear infiltrates in lung tissue (TBBx: transbronchial biopsies) and is graded by the severity of inflammation both in the vascular compartment (A0 to A4) and airway (B0 to B2R). Surveillance bronchoscopy with transbronchial biopsy was performed by physicians at 4–6 weeks, and then 3-, 6-, 9-, and 12-months post-Tx. Subsequent and/or additional bronchoscopies were performed as clinically needed. Blood samples were collected once weekly for the first 12 weeks, and subsequently at least once per month if available. Spirometry and chest radiographs were performed once a week for the first 12 weeks and subsequently at least once a month.

Initial immunosuppression for all LuTx patients consisted of induction therapy with basiliximab administered on post-operative day (POD) 0 and POD4 (each dose: 20 mg IV infused over 30 minutes), as well as methylprednisolone (1000 mg IV on POD0). Pre-operatively, tacrolimus was administered (2 mg PO x 1 for patient weight ≥ 50 kg, 1 mg PO x 1 for patient weight < 50 kg), and mycophenolate mofetil was administered (1000 mg PO x 1). LuTx patients followed a post-Tx maintenance regimen that included a calcineurin inhibitor (tacrolimus), antiproliferative agents (mycophenolate mofetil), and corticosteroids (methylprednisolone and prednisone).

Tacrolimus was continued post-operatively and administered every 12 hours, adjusted to aim for a target trough level of 12–15 ng/mL during the first year post-Tx, and 10–15 ng/mL during subsequent years post-Tx. Methylprednisolone was also continued, with one 125 mg dose every 8 hours for 24 hours on POD1, followed by 0.5 mg/kg IV daily that was gradually tapered to 10 mg daily by POD90, and then ideally further tapered to 5–10 mg daily after 1–2 years post-Tx. Methylprednisolone was changed to oral prednisone (1:1) if the patient could tolerate oral medication. Mycophenolate mofetil was continued at 1000 mg PO/IV every 12 hours. In instances of allograft rejection, immunosuppression was tailored and administered based on the severity of rejection.

### Bronchoalveolar lavage (BAL) cell isolations

BAL cells were isolated from surveillance bronchoscopy specimens or pre-Tx donor or recipient BAL samples when available. Specimens were centrifuged at 515g for 5 minutes at 4°C, the supernatant was then passed through a 40 μm filter. The filtrate was frozen at -80°C in a maximum of five 1mL aliquots, and the cell pellet was resuspended in 2mL of ACK lysis buffer, incubated for 3 minutes at room temperature, then centrifuged at 515g for 5 minutes at 4°C. The supernatant was discarded, and the cell pellet was resuspended in PBS, where it then passed through another 40 μm filter, and then cell counting (focused on lymphocyte-looking cells) was performed on the filtrate using a hemocytometer under the microscope (BH2, Olympus). In rare cases, bronchoalveolar washing (BW) fluid was collected and the cell isolation was similar as to processing BAL cells except the supernatant was not collected after the initial spin and filtering process.

### PBMC isolations

PBMCs were isolated from patient whole blood samples or pre-Tx donors or recipient whole blood samples when available. To isolate PBMCs, density gradient centrifugation was utilized. First, patient blood specimen was diluted in PBS at a 1:4 to 1:10 volume ratio. Histopaque was then added to fresh conical tubes, and the diluted blood was carefully layered on top of the Histopaque, keeping a clear separation between both layers. The volume ratio of Histopaque to dilute blood in each tube followed a ratio of 1:2. The tubes were then centrifuged at 400g for 30 minutes at 22°C, with acceleration set to 3 and brakes set to 0. The buffy coat was then carefully isolated, washed with PBS, treated with ACK lysis buffer, reconstituted with PBS, and then cell counting was performed using a hemocytometer under the microscope.

### Lymphocyte isolation from pre-Tx donor and recipient lung-associated lymph nodes (LLNs)

The surgeon removed LLNs from the explanted lung(s), and lymphocytes were isolated from pre-Tx donor or recipient samples when available. Using forceps and scissors, samples were cut into small pieces (about 5 mm^2^) and incubated for 1 hour at 37°C in culture medium (RPMI 1640, 1 mg/mL Collagenase D, and 100 IU/mL penicillin-streptomycin). After incubation, the samples were filtered through a 100μm filter and resuspended in MLR medium (AIM-V media mixed with 5% AB heat-inactivated human serum, 0.01M Hepes, and 50 μm 2-mercaptoethanol) then centrifuged for 5 minutes at 515g. If the cell pellet appeared to contain red blood cells, it resuspended in 5 mL of ACK lysis buffer & incubated for 5 minutes on ice, then brought to 50 mL with 1x RPMI, then centrifuged again. The cell pellet was then resuspended in MLR media, passed through a 40 μm filter, and counted using a hemocytometer under the microscope (BH2, Olympus).

### Lymphocyte isolation from pre-Tx lung explant tissues

Cells were isolated from patient pre-Tx lung explant tissues when available. Using forceps and scissors, lung tissues were cut into 4–10 mm^3^ pieces and then treated for 1.5 hours at 37°C in sterile-filtered isolate buffer (RPMI 1640, 1 mg/mL Collagenase D, 100 IU/mL penicillin-streptomycin, 10 μg/mL DNAse, and 1 μg/mL Amphotericin B). Residual tissue pieces were further broken down using the gentleMACS Dissociator, and the supernatant was filtered. The filtrate was centrifuged at 450g for 10 minutes at 21°C and the cell pellet was resuspended in MLR medium (AIM-V media mixed with 5% AB heat-inactivated human serum, 0.01M Hepes, and 50 μm 2-mercaptoethanol). This resuspension was then mixed with an 88% isotonic Percoll solution (9:1:1.36 ratio of stock Percoll, 10X PBS, and 1X HBSS without Ca^2+^ respectively) at a 1:1 volume ratio. Lung tissue cells were then isolated from this mixture via density gradient centrifugation for 20 minutes at 800g for 20 minutes, with the brakes set to 0. The supernatant was aspirated, and the remaining cell pellet was treated with ACK lysis buffer before being washed, filtered, and counted using a hemocytometer under the microscope (BH2, Olympus).

### Human leukocyte antigen (HLA) allotype-specific staining and cellular staining by flow cytometry

Based on the clinically available HLA typing for each LuTx donor and recipient (Table S2), various monoclonal HLA class I allotype-specific antibodies were selected and then assessed for their ability to discriminate between donor and pre-Tx recipient cells as we published previously [26]. During this quality control process, each HLA allotype-specific antibody was used in conjunction with pan-HLA-ABC antibody, and those that adequately discriminated between donor and pre-Tx recipient PBMCs were included when tailoring lineage-specific antibody panels for each LuTx patient (Table S2). Cellular staining was performed on up to 0.5M PBMCs or BAL cells from the patient sample and compared to healthy control PBMCs isolated from the blood product provided by the New York Blood Bank. Staining was performed in accordance with previous published protocol [26]. Briefly, 1mL of FACS buffer was added to each sample and then centrifuged at 465 g for 5 minutes at 4°C. The supernatant was poured off, then 2.5 µL of Fc block followed by 5 µL of each antibody in the patient’s lineage-specific panel was added to each tube as the primary staining. Multiple flow cytometry antibodies were used in this process (Table S3). Following a 30-minute incubation at 4°C, the sample was centrifuged again after 1mL of FACS buffer was added, then the supernatant was poured off and cells were resuspended at 10x10^6^ cells/mL of FACS buffer before 5 µL of the secondary antibody was added and incubated at 4°C for 30 minutes. The samples were then centrifuged with 1 mL of FACS buffer, the supernatant was later poured off, and cells were resuspended in FACS buffer before the final step where 2.5 µL of DAPI (2ug/ml stock) was added. Cells were kept at 4°C (for a maximum of 12 hours) before acquisition on either the BD LSR II flow cytometer or the Cytek Aurora flow cytometer. Data analysis was performed using FlowJo software (V10.10.0, BD Biosciences).

### Carboxyfluorescein succinimidyl ester (CFSE)-mixed lymphocyte reactions (MLRs) and cell sorting

LuTx MLR assays were performed as described previously [18, 53]. GvH and HvG MLRs were set up with thawed pre-Tx donor and pre-Tx recipient LLN cells. In each MLR, 200,000 CFSE-labeled responders and 200,000 violet-dye-labeled irradiated (35 Gy, X-RAD320) stimulators in MLR medium (AIM-V media mixed with 5% AB heat-inactivated human serum, 0.01M Hepes, and 50 μm 2-mercaptoethanol) were plated together in wells of a round-bottom 96-well plate.

After incubation at 37°C for six days, the cultures were harvested, and the cells were stained with an antibody panel including DAPI, CD45 V500, CD3 PerCP-Cy5.5, γδTCR PE-Cy7, CD4 redfluor 710 and CD8 APC-Cy7. Then, the cells underwent FACS sorting on a BD Influx cell sorter to isolate four violet-dye-negative, DAPI negative, CD45 positive cell populations: CD3^+^γδTCR^-^CD4^+^CFSE^low^ and CD3^+^γδTCR^-^CD8^+^CFSE^low^, which represent the proliferated CD4^+^ and CD8^+^ T cells, and CD3^+^γδTCR^+^CFSE^low^, and CD3^+^γδTCR^+^CFSE^high^, which represent the proliferated and non-proliferated γδTCR T cells for other projects.

### DNA extraction and TCRβ CDR3 DNA sequencing

Genomic DNA was isolated from different tissue types from LuTx patients and their donors using the QIAGEN DNeasy Blood and Tissue Kit. FFPE tissue curls of post-Tx lung bronchoscopy specimens were requested from the Pathology Core at the Columbia University Irving Medical Center in order to extract genomic DNA, and the QIAGEN DNA FFPE Tissue Kit was used to isolate the DNA. Purity and yield of all DNA samples were assessed on the Nanodrop (Thermo Fisher Scientific). The DNA was frozen at -20°C and shipped on dry ice to Adaptive Biotechnologies for high-throughput TCRβ sequencing. Samples for TCRβ sequencing were summarized in Fig. S2.

### TCRβ CDR3 data processing and analysis

TCR sequencing data was retrieved from Adaptive Biotechnology’s ImmunoSEQ Annalyzer. Data analysis followed our previously published method [19, 21, 23]. Briefly, CD8 versus CD4 sorting error was corrected by removing sequences detected in both populations at high to low frequency ratio less than 5:1. Donor- and recipient-shared CDR3s at the nucleotide level were removed due to not being clearly assigned to either origin. Separate CD4 and CD8 tables containing clonal frequencies in pre-Tx unstimulated samples, CFSE^low^ stimulated cells, and post-Tx samples were compiled and renormalized. Alloreactive clones were defined by 2-fold or greater expansion in stimulated compared with unstimulated pre-Tx cells by minimum frequency of 0.002% in CFSE^low^ populations when using template counts, which serves to ensure 85% repeatability, as determined by power analysis. Mappable clones refer to clones that were detectable in sequenced pre-Tx lung, lymph node, and/or MLR CFSE^low^ populations from the donor or recipients. Cumulative frequency was calculated as a percentage of all sequences weighted by copy numbers in designated populations (Table S4). The unique clone numbers of mappable clones, mappable alloreactive clones, and the cumulative frequency of alloreactive clone of post-Tx samples were summarized in Table S5. The number of pathogen-reactive TCRs collected from public available database were listed in Table S6 [27–32].

## Software and Statistical Analysis

R (R-4.4.3) and RStudio (2024.12.1) were used to analyze TCR-seq data and generate circle plots. GraphPad Prism (v 10.4.2, GraphPad Software) was used to produce figures. A Shapiro– Wilk test was performed to determine normality of data distributions. For the two unpaired data sets that meet the assumption of a normal distribution, a parametric unpaired t test was performed. For the two paired data sets that meet the assumption of a normal distribution, a parametric paired t test was performed. For the two paired data sets that didn’t meet the assumption of a normal distribution, a non-parametric Wilcoxon test was performed to compare the median ranks. A non-parametric Kruskal–Wallis test followed by Dunn’s multiple comparisons test was performed for comparisons among three or more unpaired independent groups that didn’t meet the assumption of a normal distribution, to compare the median ranks. For three or more independent groups with paired data that didn’t follow the assumption of a normal distribution, a non-parametric Friedman test followed by Dunn’s multiple comparisons test was performed.

## Data and materials availability

Raw TCRβ-seq data are available at https://clients.adaptivebiotech.com and a review account will be available for peer review. The R code used to analyze TCRβ-seq data is available in the GitHub repository at https://github.com/drjiaowy/LuTx_manuscript.git.

## Data Availability

All data produced in the present study are available upon reasonable request to the authors.

## Acknowledgements

We thank the Flow Cytometry Core and Human Studies Core at Columbia Center for Translational Immunology (CCTI) for their excellent services. We gratefully acknowledge the generosity of the donor families, our LuTx patients and their families for making this study possible.

## Funding

The study is supported by the Thomas Kully Immunology Fund and the Nelson Family Transplant Innovation Award Program at the Columbia University Irving Medical Center (to J.F.). W.J. was supported by the Nelson Family Transplant Fellowship and Innovation Award at Columbia University Irving Medical Center. Research reported here was performed in the CCTI Flow Cytometry Core, supported in part by the Office of the Director, National Institutes of Health (NIH) awards S10OD030282 and S10OD020056.

## Author contributions

J.F. designed the study. W.J., K.D.L., T.Y., J.H.W. and J.F. performed the experiments. C.B.M., A.P.R., M.K., V.M., K.R., A.V., J.C., L.B., J.S., P.L., F.D., S.A. and J.F. coordinated the clinical sample and data collection. J.S., P.L. and F.D. performed the lung transplantation. L.B. and S.A. performed the routine patient care. W.J., K.D.L., T.Y., J.H.W. and J.F. performed data analysis. W.J. and J.F. wrote the codes to identify and track alloreactive and microbial reactive TCR clones. W.J. and J.F. wrote the final report. All authors contributed to the editing of the final report. All authors agree to all the content of the submitted manuscript.

## Declaration of interests

J.F. served as a Scientific Consultant for Adaptive Biotechnologies from June 2022 to May 2023.

## Privacy Statement

Identification numbers for patients and deceased organ donors are not known to anyone outside the research group.

---

**Fig. S1.**
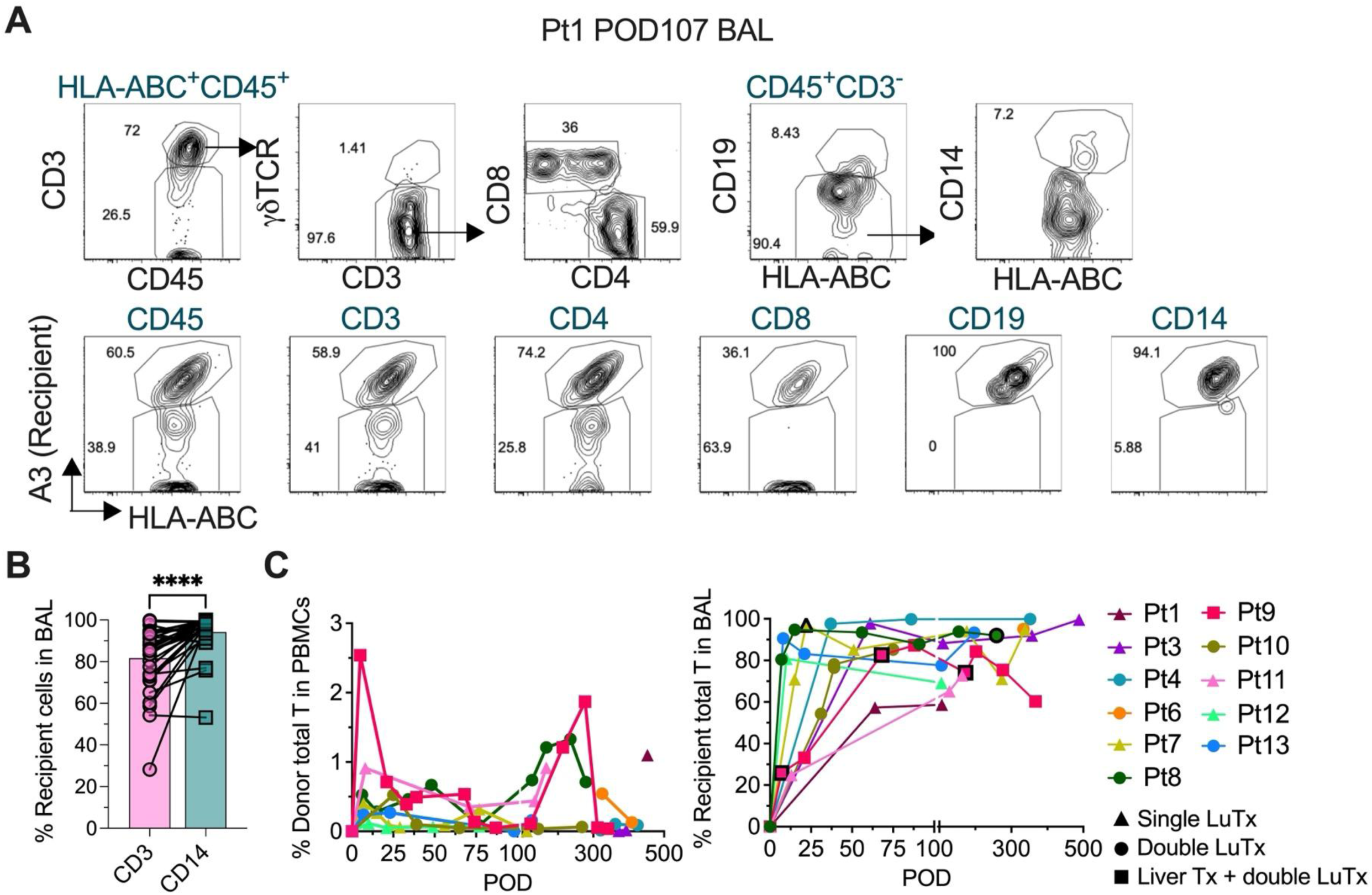
Immune cell chimerism analysis in post-Tx samples. (A) Representative gating strategy for T/B cell/monocyte chimerism in post-Tx BAL (Pt1 POD107). Recipient cells were identified by HLA-A3+ HLA-ABC+ expression. Donor cells were identified by HLA-A3- HLA-ABC+ expression. (B) Recipient-derived CD3+ T cells and CD14+ monocytes in BAL samples across all patients (POD7–352). (C) Longitudinal chimerism patterns showing donor T cell percentages in PBMC (left panel) and recipient T cell percentages in BAL (right panel) from Pt1 to Pt13. Black-outlines symbols indicate timepoints with pathology-determined ACR. Wilcoxon test was performed for panel B. ****p < 0.0001.

**Fig. S2.**
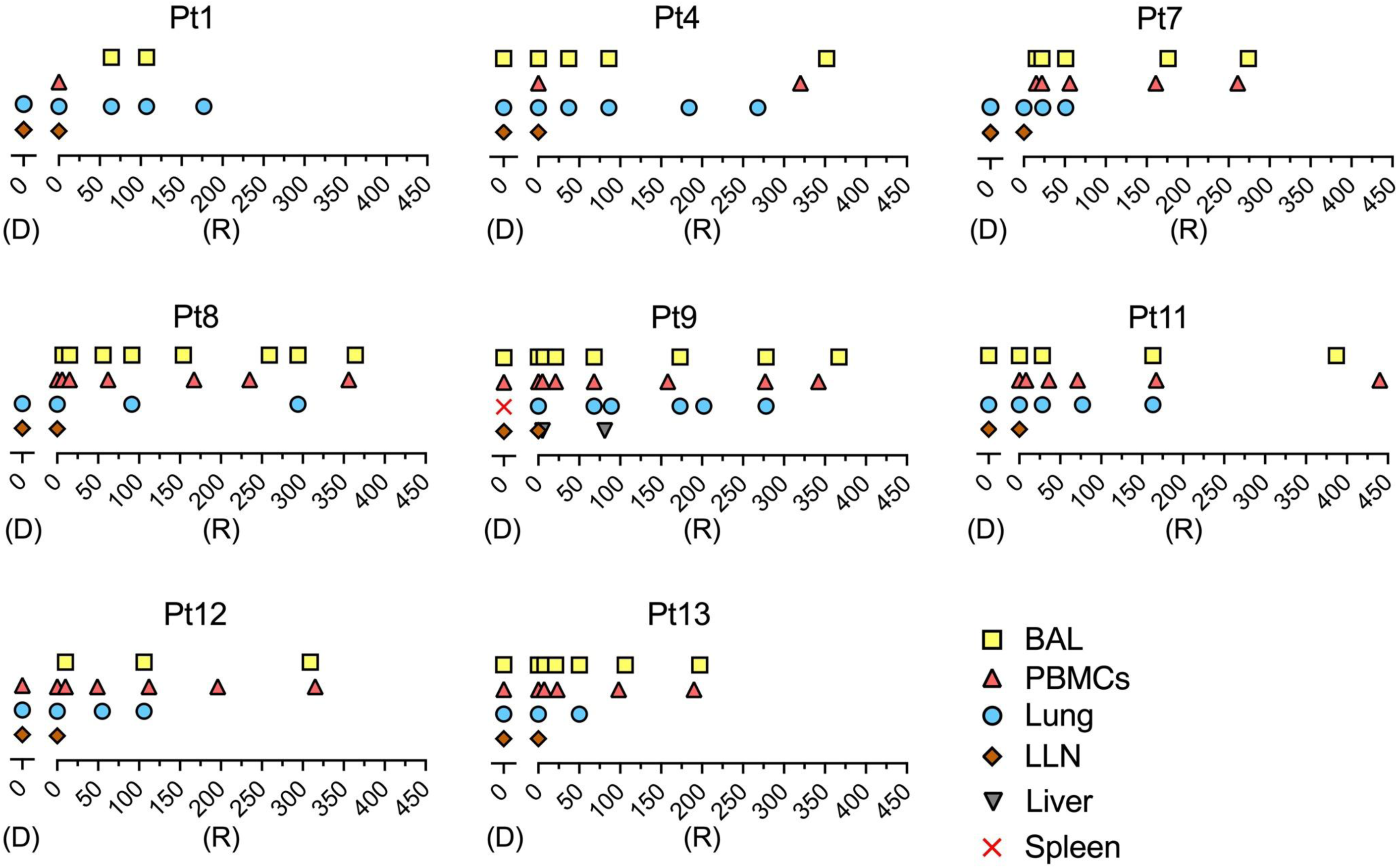
Sample collection overview for TCRβ sequencing analysis. Summary of all pre-Tx and post-Tx specimens with available TCR-β sequencing data. Samples are annotated by origin (D: donor; R: recipient) and tissue types.

**Fig. S3.**
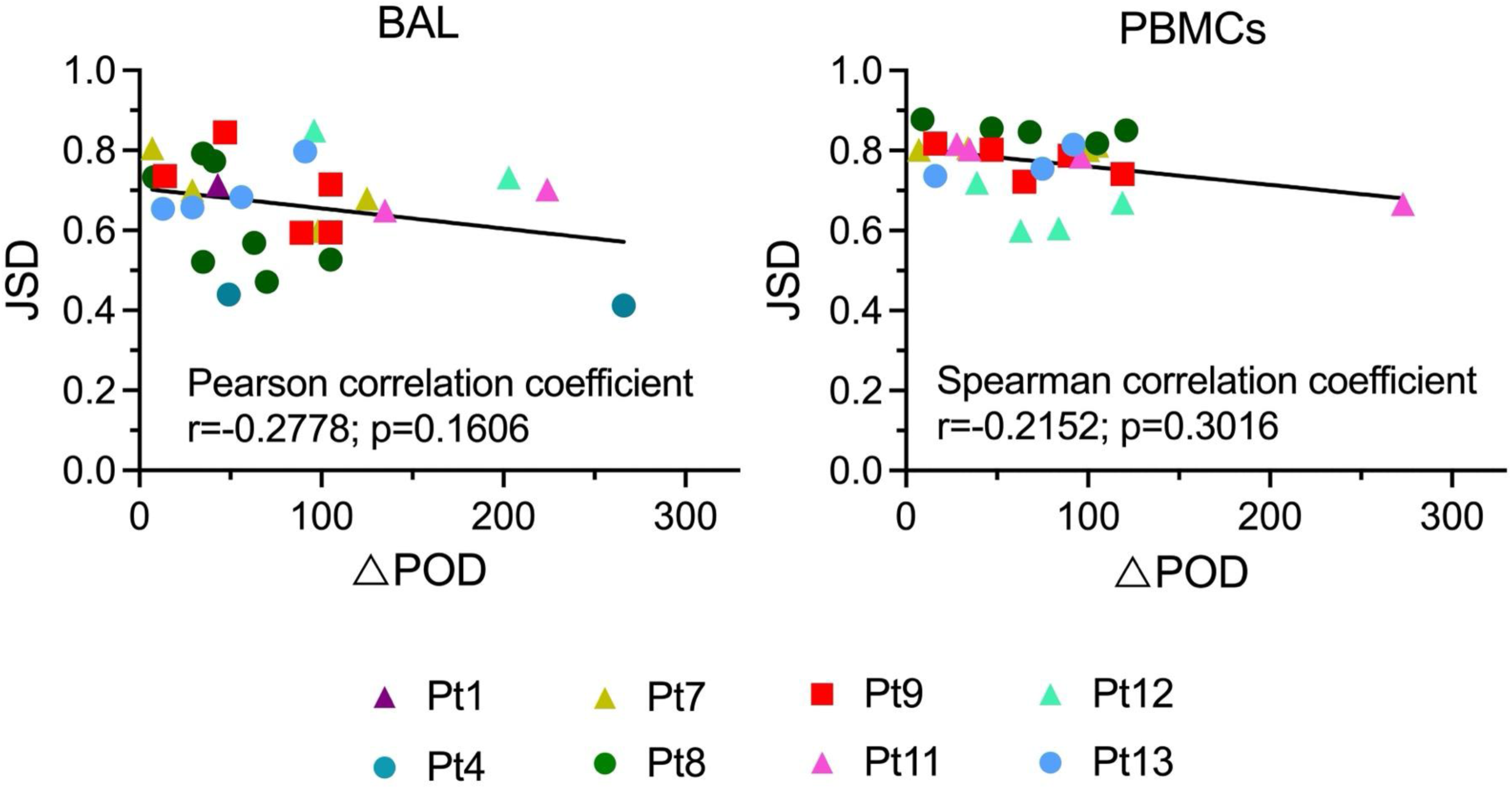
TCR repertoire stability analysis using Jensen-Shannon Divergence (JSD). JSD values are plotted against the time interval (△POD, in days) between consecutive sampling time points. Correlation analyses were performed using Pearson’s correlation for normally distributed data and Spearman’s rank correlation for non-normally distributed data.

**Fig. S4.**
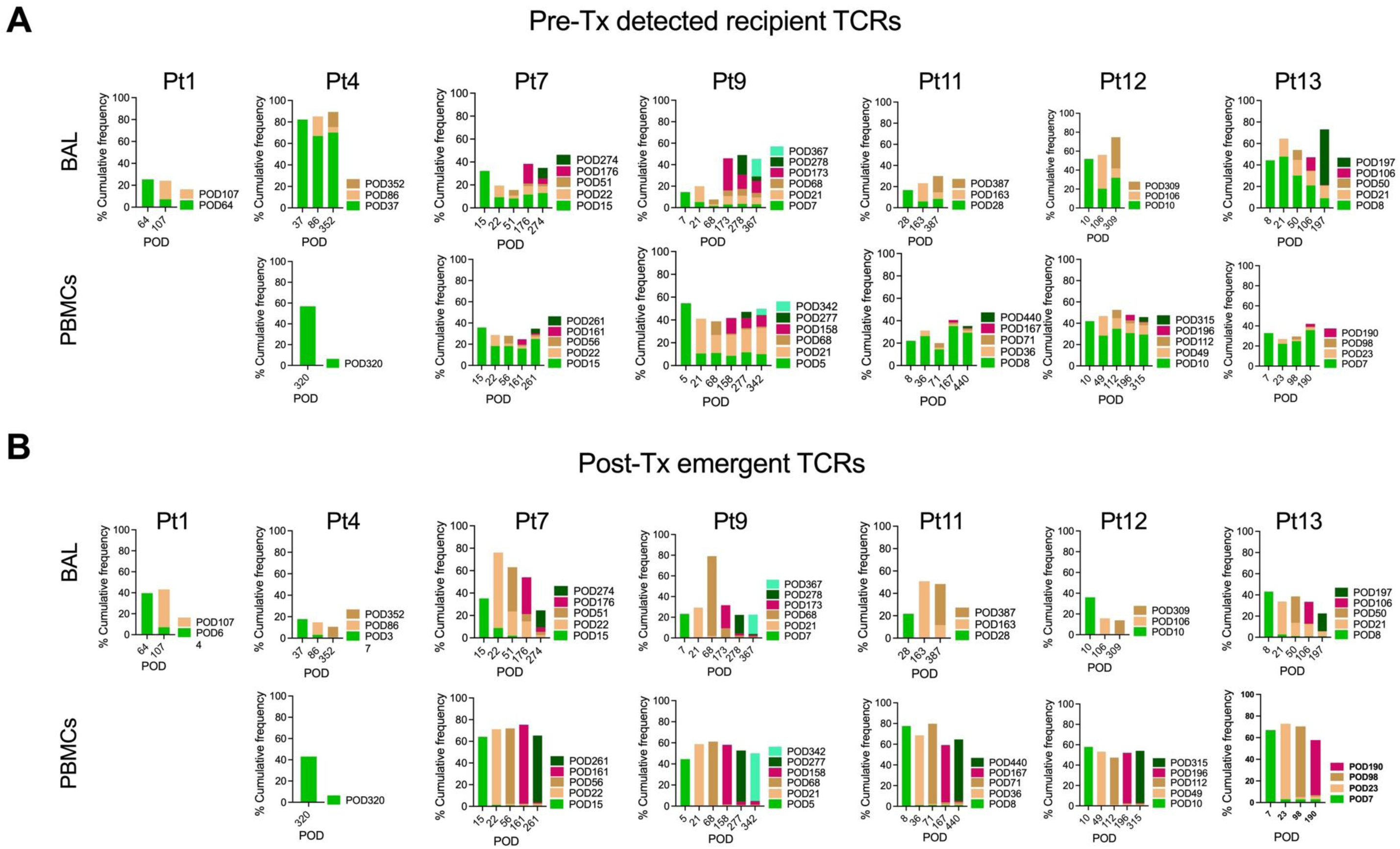
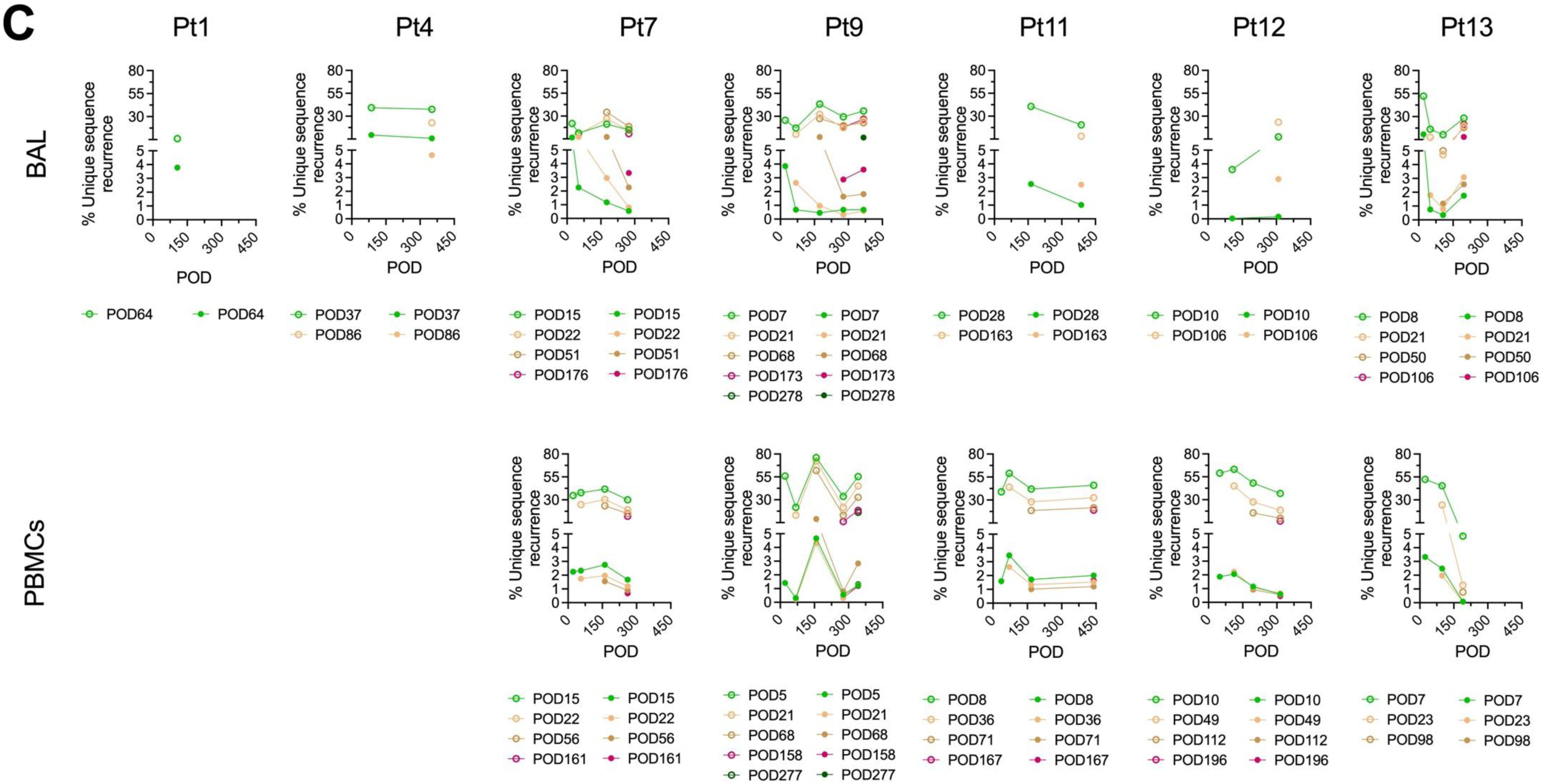
Contribution of pre-existing and newly emergent TCRs to post-Tx repertoire maintenance. Pre-Tx detected recipient TCRs (A) and post-Tx newly detected TCRs (B) both contribute to TCR repertoire diversity in BAL and PBMCs. (C) Pre-Tx detected recipient TCRs (open symbols) and post-Tx newly detected TCRs (solid symbols) demonstrate distinct recurrence patterns during immune reconstitution.

**Fig. S5.**
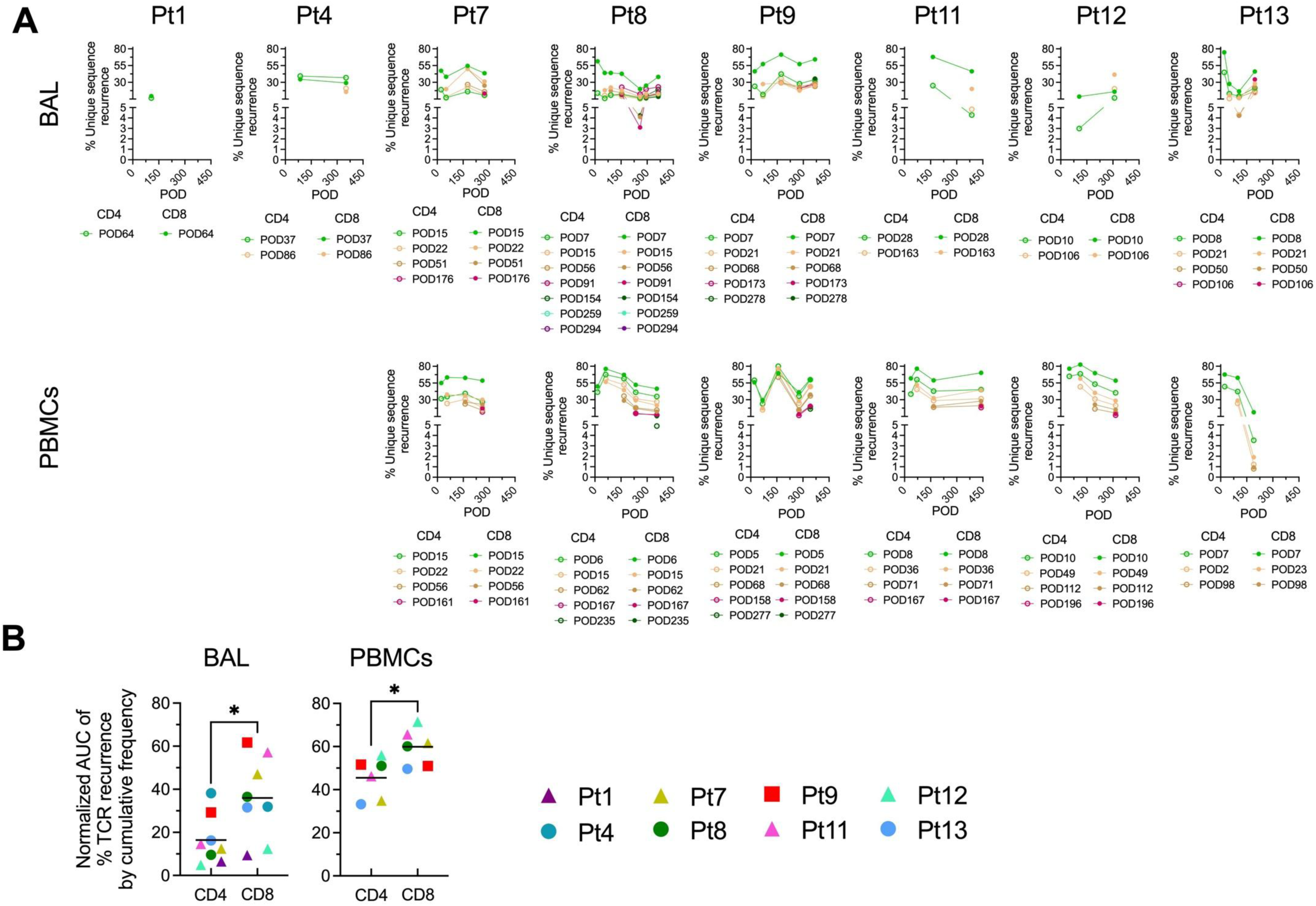
Recurrence dynamics of pre-existing recipient TCR subsets. (A) Recurrence proportions of pre-Tx detected recipient TCRs stratified by CD4+ and CD8+ subsets in BAL and blood samples. (B) Comparison of recurrence between the earliest detected pre-Tx recipient CD4+ and CD8+ TCR subsets across patients. Recurrence values were normalized by the area under curve (AUC) divided by the observation time span (as shown in panel A). Paired t test was performed for panel B. Solid horizontal bars indicate means. *p < 0.05.

**Fig. S6.**
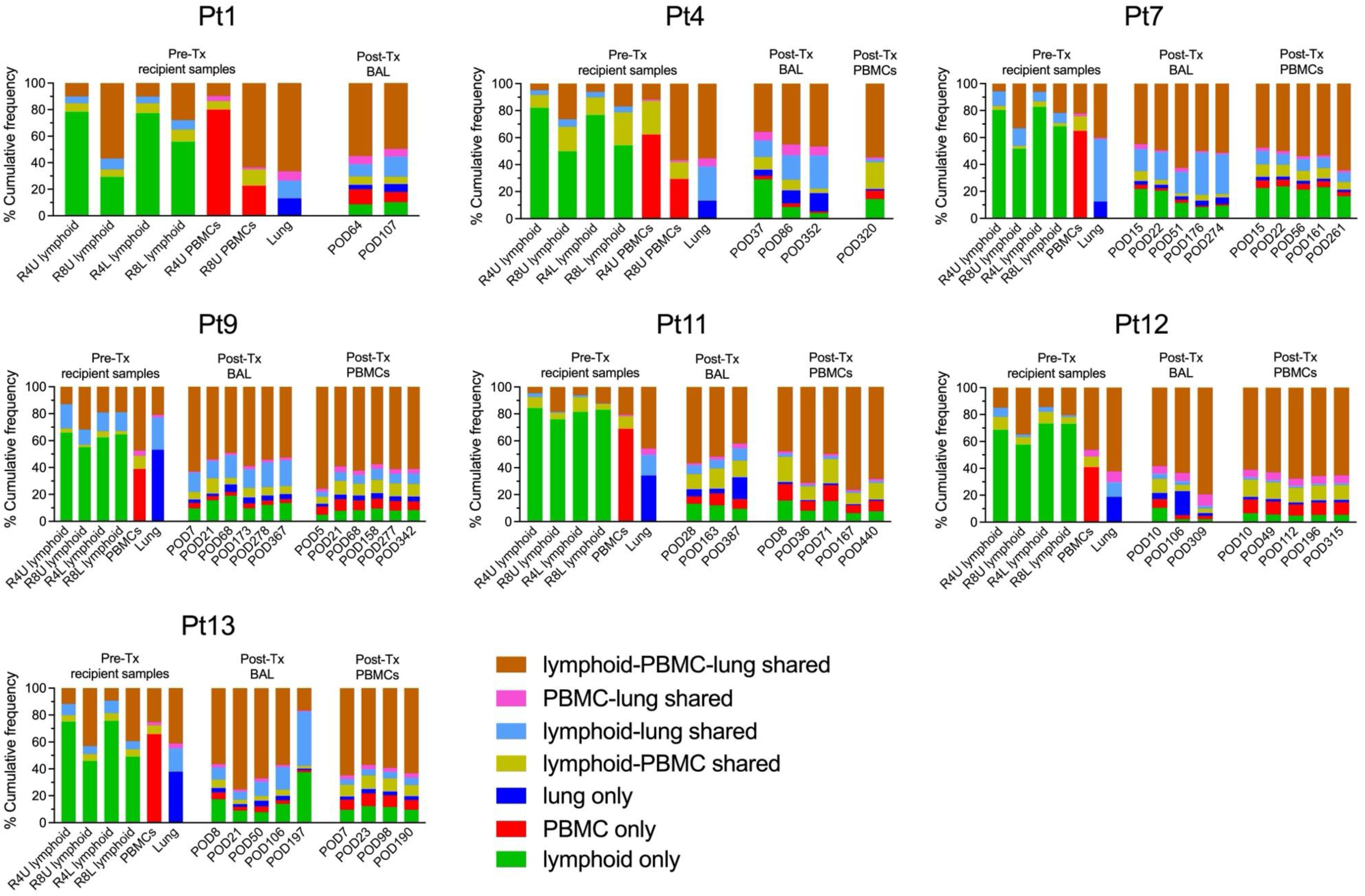
Composition of T cell subsets by tissue origin. Cumulative frequencies of recipient TCRs classified as “lymphoid-PBMC-lung shared”, “lymphoid-lung shared”, “PBMC-lung shared”, “lymphoid-PBMC shared”, “lymphoid only”, “PBMC only”, and “lung only” in pre-Tx and post-Tx samples. R4U: recipient CD4 unstimulated; R8U: recipient CD8 unstimulated; R4L: recipient CD4 CFSE^low^; R8L: recipient CD8 CFSE^low^.

**Fig. S7.**
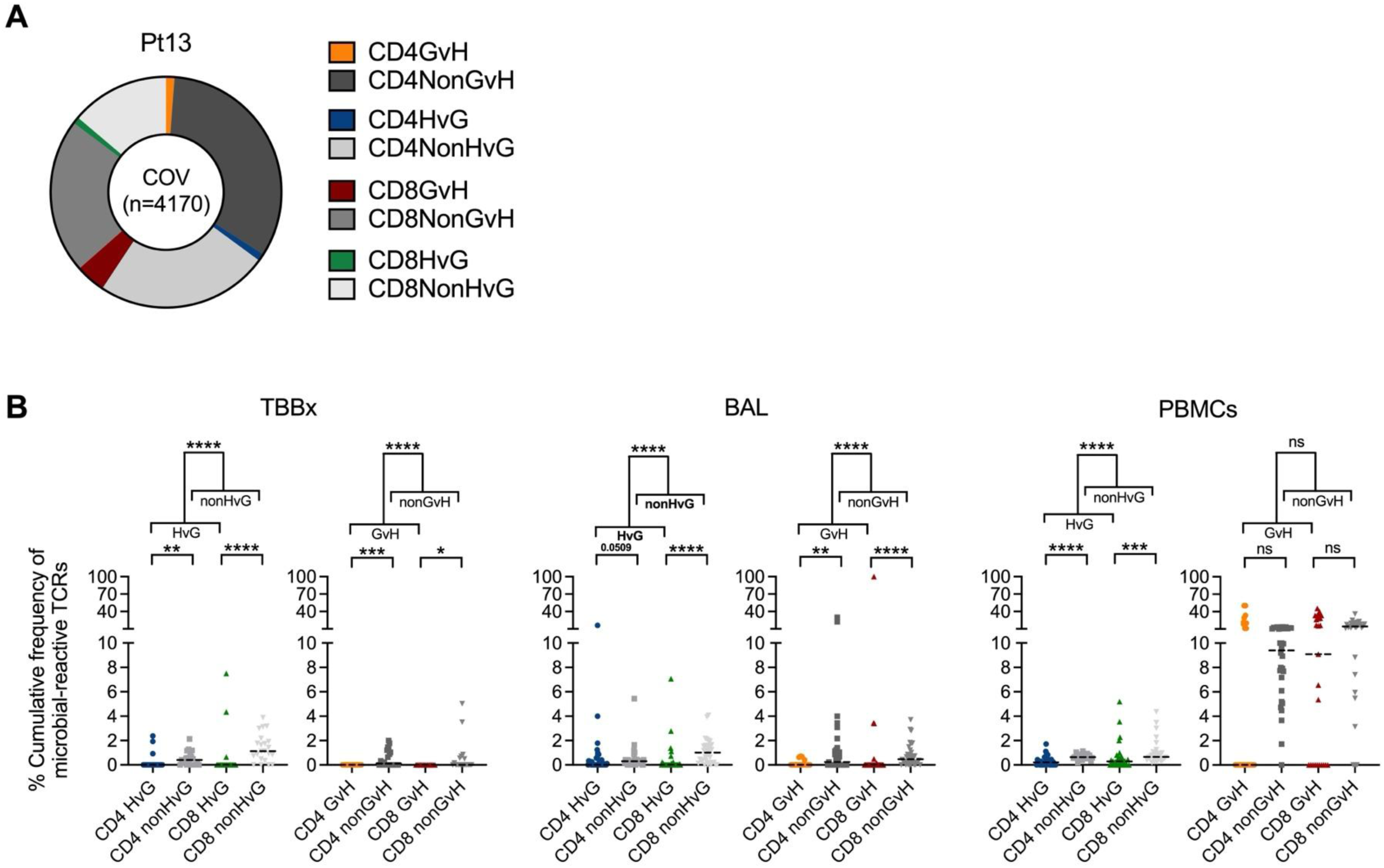
Characterization of COVID-19-reactive TCRs by alloreactivity status. (A) Composition of COVID-19-specific TCRs in Pt13 at the unique sequence level subgrouped by alloreactive vs nonalloreactive identification in pre-Tx MLRs, including CD4 and CD8 GvH and nonGvH, HvG and nonHvG categories. Pie chart centers indicate total unique TCRs reactive to COVID-19 in all samples sequenced from Pt13. (B) Cumulative frequency of COVID-19-specific TCRs in post-Tx samples (n=8 patients), stratified by alloreactive (CD4 HvG, CD8 HvG, CD4 GvH, CD8 GvH) vs nonalloractive (CD4 nonHvG, CD8 nonHvG, CD4 nonGvH, CD8 nonGvH) identification. Wilcoxon tests were performed for panel B. Dotted horizontal bars indicate medians. *p < 0.05, **p < 0.01, ***p < 0.001, ****p < 0.0001.)

**Fig. S8.**
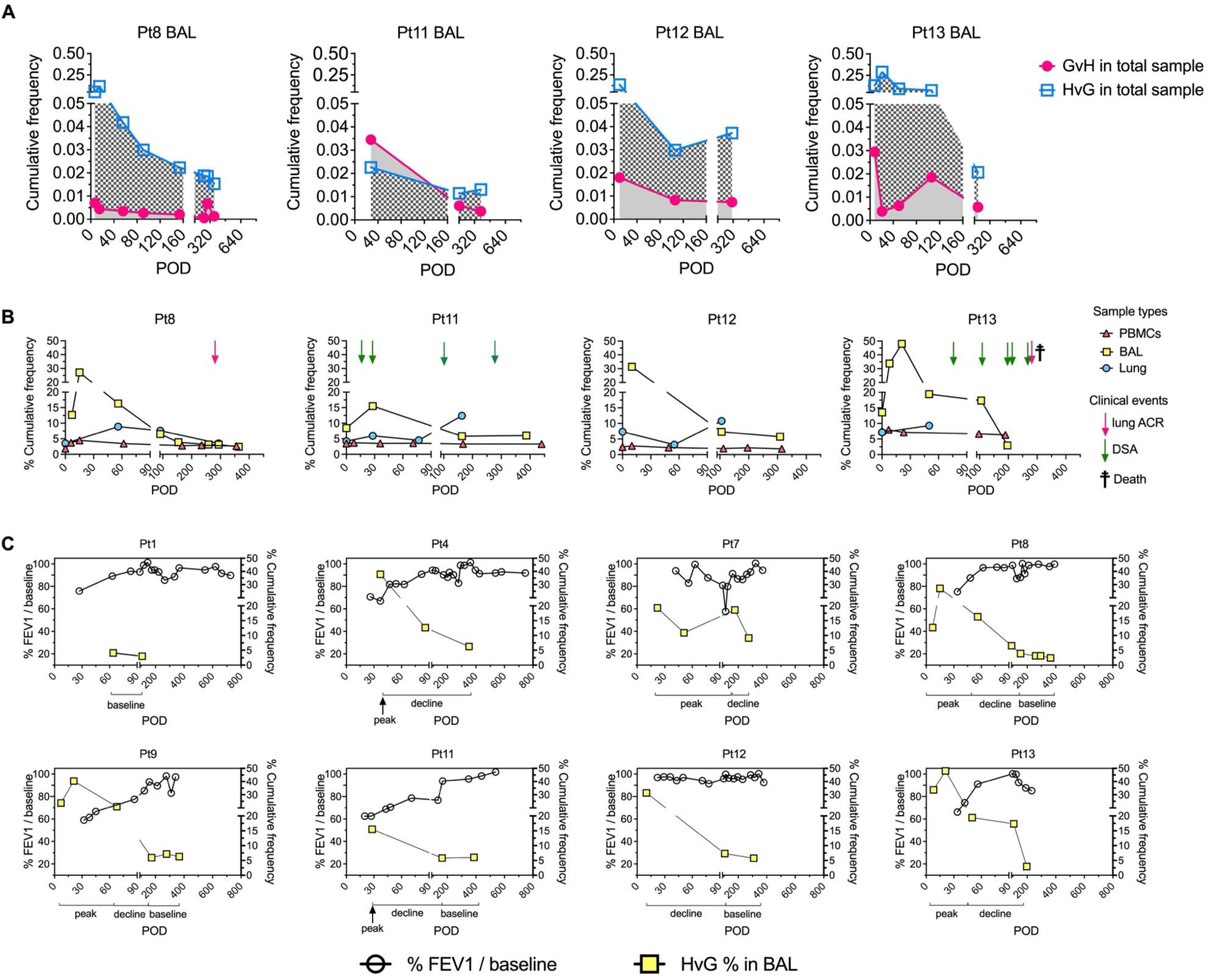
Tracking of alloreactive clones and pulmonary function post-Tx. (A) Cumulative frequency of GvH and HvG clones in BAL specimens in Pts8, 11, 12, and 13. (B) Percentage cumulative frequency of HvG-reactive sequences among recipient mappable repertoires in post-Tx PBMC, BAL and lung allografts in rPts8, 11, 12, and 13. (C) Left Y axis: The FEV1/baseline FEV1 proportion cross time curve of all the patients. Right Y axis: Percentage cumulative frequency of HvG-reactive sequences among recipient mappable repertoires in post-Tx BAL labeled with peak, decline and baseline phases. Some peak phases were defined as a range rather than a single peak value for HvG clones in BAL, due to the lack of FEV1/baseline data corresponding to the peak HvG values. Additionally, the earliest HvG mesurements in BAL were excluded from the correlation analysis in Fig. 5D because of missing adjacent FEV1/baseline data. FEV1: forced expiratory volume in one second.

**Table S1.** Epidemiological and clinical characteristics of patients. F: female. M: male. COVID-19: coronavirus disease 2019. CMV: Cytomegalovirus. BKV: BK polyomavirus. EBV: Epstein-Barr virus. HBV: Hepatitis B virus. HCV: Hepatitis C virus. VZV: Varicella zoster virus. Precise ages were replaced with age ranges as required.

| Pt # | Recipient Age at Tx (yrs) | Recipient Sex | Indication for Transplant | Type of LuTx | Donor Age (yrs) | Donor Sex | Infection | Rejection | CLAD | Death (Cause of death) |
| --- | --- | --- | --- | --- | --- | --- | --- | --- | --- | --- |
| 1 | 61-65 | F | Pulmonary emphysema | Single | 46-50 | M | Kiebsiella /COVID-19 /CMV | No | No | No |
| 2 | 71-75 | M | Chronic atrial fibrillation | Single | 46-50 | M | No | No | No | Yes (POD166: cardiopulmonary arrest secondary to complications related to hypoxia and atrial fibrillation and rapid ventricular response) |
| 3 | 66-70 | M | Interstitial lung disease | Single | 11-15 | M | Stenotrophomonas maltophilia / Lomentospora | POD327: A0B1R | Yes | Yes (POD1427: respiratory failure. Contributory causes: gastrointestinal hemorrhage and skin malignancy). |
| 4 | 61-65 | F | Interstitial lung disease, pulmonary hypertension | Double | 16-20 | F | aspergillus / pseudomonas aeruginosa / klebsiella / mycobacterium avium / CMV / BKV | POD924: minimal ACR (A1B0) | Yes | Yes (POD1556: respiratory failure. Contributory cause: liver failure). |
| 5 | 61-65 | F | Pulmonary hypertension | Double | 61-65 | M | No | No | No | Yes (POD16: bowel perforation with multisystem organ failure) |
| 6 | 66-70 | M | Pulmonary hypertension | Double | 21-25 | M | Stenotrophomonas maltophilia / CMV | No | No | No |
| 7 | 66-70 | M | Interstitial lung disease, chronic obstructive pulmonary disease, idiopathic pulmonary fibrosis, bronchiolitis obliterans with organizing pneumonia | Single | 51-55 | F | aspergillus | POD23: mild ACR (A2B0) | No | No |
| 8 | 51-55 | F | Idiopathic pulmonary fibrosis, Sjogren's related nonspecific interstitial pneumonitis, severe pulmonary hypertension | Double | 31-35 | F | CMV / pluralibacter | POD259: minima I ACR (A1B0) | No | No |
| 9 | 26-30 | M | Cystic fibrosis | Double lung + Liver | 31-35 | F | scedosporium apiospermum / enterobacter cloacae / staph aureus / EBV | POD4: mild-moderate ACR in liver; POD67: mild ACR in lung (A2B1R); POD69: mild ACR in liver; POD80: minimal ACR in liver; POD172: minimal ACR in lung (A1BX) | No | No |
| 10 | 41-45 | F | Pulmonary hypertension | Double lung | 36-40 | F | Candida tropicalis / rhino / enterovirus / CMV | No | No | No |
| 11 | 31-35 | M | Interstitial lung disease related to connective tissue disease, systemic sclerosis | Single Right | 31-35 | M | staph aureus / CMV / E. coli / rhizopus spp / coronavirus HKU1 | No | No | No |
| 12 | 61-65 | F | Interstitial lung disease, idiopathic pulmonary fibrosis | Single Right | 46-50 | F | HBV / HCV / penicillium / parainfluenza virus 1 / actinomyces | No | No | No |
| 13 | 31-35 | M | Pulmonary sarcoidosis | Double | 11-15 | M | CMV / EBV / VZV / measles / rubella / candida / TB / pseudomonas aeruginosa / mycobacterium / parainfluenza | POD279: mild ACR (A2BX) | No | Yes (POD303: multisystem organ failure) |

**Table S2.** HLA-A, B typing and anti-HLA allotype antibodies used to distinguish donor from recipient cells in lung transplant recipients. HLA-A09 is a broad antigen HLA-A serotype that recognized the HLA-A23 and HLA-A24 serotypes. HLA-A28 is a broad antigen HLA-A serotype that recognized the HLA-A68 and HLA-A69 serotypes. HLA-B12 is a broad antigen HLA-B serotype that recognized the HLA-B44 and HLA-B45 serotypes.

| Pt# | Recipient HLA-A, B type | Donor HLA-A, B type | HLA allotype-specific antibodies used to distinguish between recipient and donor cells |
| --- | --- | --- | --- |
| 1 | **A03** / A29 | A30 / A36 | Anti-HLA A3 APC |
|  | B27 / B44 | B42 / B53 |  |
| 2 | **A03** / A29 | A30 / A36 | Anti-HLA A3 APC, Anti-HLA B12 FITC |
|  | B61 / **B44** | B42 / B53 |  |
| 3 | **A02** / A03 | A01 / A03 | Anti-HLA A2/A28 biotin, Anti-HLA B8 FITC |
|  | B39 / B44 | **B08** / B35 |  |
| 4 | **A03** / A31 | A24 / **A68** | Anti-HLA A3 APC, Anti-HLA A2/A28 biotin |
|  | B35 / B58 | B71 / B35 |  |
| 5 | A02 / **A03** | A02 / A29 | Anti-HLA A3 APC, Anti-HLA B12 FITC |
|  | B35 / B61 | B35 / **B44** |  |
| 6 | A34 / A74 | **A02** / A24 | Anti-HLA A2/A28 biotin |
|  | B72 / B53 | B35 / B61 |  |
| 7 | A02 / A32 | A02 / **A03** | Anti-HLA B12 FITC, Anti-HLA A3 APC |
|  | **B44** / B52 | B13 / B51 |  |
| 8 | A33 / **A68** | A31 / A32 | Anti-HLA A2/A28 biotin |
|  | B07 / B53 | B35 / B51 |  |
| 9 | **A02** / **A24** | A30 / A74 | Anti-HLA A2 FITC, Anti-HLA A09 biotin |
|  | B35 / B45 | B72 / B42 |  |
| 10 | A01 / A25 | A01 / A31 | Anti-HLA B7 APC |
|  | B08 / B18 | **B07** / B08 |  |
| 11 | A01 / **A68** | A33 / A66 | Anti-HLA A2/A28 biotin |
|  | B35 / B61 | B65 / B53 |  |
| 12 | A02 / A30 | A02 / A11 | Anti-HLA B12 FITC |
|  | B71 / B53 | B35 / **B44** |  |
| 13 | A02 / **A24** | A02 / A33 | Anti-HLA A9 FITC, Anti-HLA Bw4 biotin |
|  | B60 / B57 | B39 / B65 |  |
|  | **Bw4** / Bw6 | Bw6 / Bw6 |  |

**Table S3.** List of flow cytometric antibodies used in the study.

| Antibody | Source | Identifier |
| --- | --- | --- |
| Mouse anti-Human CD3 (clone SP34-2) PerCP-Cy5.5 | BD Biosciences | Cat#552852; RRID: AB_394493 |
| Mouse anti-Human CD4 (clone OKT4) AF700 | Tonbo | Cat#80-0048; RRID: AB_2621976 |
| Mouse anti-Human CD8 (clone SK1) APC-Cy7 | BD Biosciences | Cat#561945; RRID: AB_396892 |
| Mouse anti-Human CD8 (clone SK1) Biotin | BioLegend | Cat#344720; RRID: AB_2828353 |
| Mouse anti-Human CD14 (clone M5E2) FITC | BioLegend | Cat#982502; RRID: AB_2616906 |
| Mouse anti-Human CD14 (clone M5E2) APC-Cy7 | BioLegend | Cat#301820; RRID: N/A |
| Mouse anti-Human CD19 (clone SJ25C1) BUV496 | BD Biosciences | Cat#612938; RRID: AB_2870221 |
| Mouse anti-Human CD28 (clone CD28.2) Pacific Blue | BioLegend | Cat#302928; RRID: AB_10641279 |
| Mouse anti-Human CD31 (clone WM-59) BV605 | BD Biosciences | Cat#562855; RRID: AB_2737841 |
| Mouse anti-Human CD45 (clone HI30) PE-CF594 | BD Biosciences | Cat#562279; RRID: AB_11154577 |
| Mouse anti-Human CD45RA (clone HI100) Brilliant Violet 510 | BioLegend | Cat#304142; RRID: AB_2561947 |
| Mouse anti-Human CD49a (clone SR84) BUV395 | BD Biosciences | Cat#742363; RRID: AB_2740721 |
| Mouse anti-Human CD56 (clone HCD56) Brilliant Violet 605 | BioLegend | Cat#318334; RRID: AB_2561912 |
| Mouse anti-Human CD69 (clone FN50) Brilliant Violet 650 | BioLegend | Cat#310934; RRID: AB_256315 |
| Mouse anti-Human CD103 (clone Ber-ACT8) Brilliant Violet 711 | BioLegend | Cat#350222; RRID: AB_2629651 |
| Mouse anti-Human CD197 (CCR7) (clone G043H7) PE | BioLegend | Cat#353204; RRID: AB_10913813 |
| Mouse anti-Human CD314 (NKG2D) (clone 1D11) APC | BioLegend | Cat#320808; RRID: AB_492962 |
| Mouse anti-Human HLA-A2/A28 (clone N/A) Biotin | One Lambda | Cat#BIH0037; RRID: N/A |
| Mouse anti-Human HLA-A2/A28 (clone REA142) FITC | Miltenyi Biotech | Cat#130-099-601; RRID: AB_2652040 |
| Mouse anti-Human HLA-A2 (clone BB7.2) FITC | BioLegend | Cat#343304; RRID: AB_1659245 |
| Mouse anti-Human HLA-A3 (clone GAP.A3) APC | Thermo Fisher Scientific | Cat#17-5754-42; RRID: AB_2573220 |
| Mouse anti-Human HLA-A9 (clone N/A) Biotin | One Lambda | Cat#BIH0964; RRID: N/A |
| Mouse anti-Human HLA-A9 FITC (clone N/A) | One Lambda | Cat#FH0964; RRID: N/A |
| Mouse anti-Human HLA-A29 (clone N/A) Biotin | One Lambda | Cat#BIH0155; RRID: N/A |
| Mouse anti-Human HLA-ABC (clone G56-2.6) APC | BD Biosciences | Cat#555555; RRID: AB_398603 |
| Mouse anti-Human HLA-ABC (clone G46-2.6) BV786 | BD Biosciences | Cat#740982; RRID: AB_2740606 |
| Mouse anti-Human HLA-B8 (clone N/A) FITC | One Lambda | Cat#FH0536A; RRID: N/A |
| Mouse anti-Human HLA-B12 (clone REA138) FITC | Miltenyi Biotech | Cat#130-099-860; RRID: AB_2652098 |
| Mouse anti-Human HLA-Bw4 (clone REA274) Biotin | Miltenyi Biotech | Cat#130-123-859; RRID: AB_2889687 |
| Mouse anti-Human HLA-Bw6 (clone REA143) APC | Miltenyi Biotech | Cat#130-099-845; RRID: AB_2652026 |
| Streptavidin Alexa Fluor 594 | Thermo Fisher Scientific | Cat#S32356; RRID: N/A |
| Streptavidin BUV737 | BD Biosciences | Cat#564293; RRID: AB_2869560 |
| Mouse anti-Human gdTCR (clone IMMU510) PE-Cy7 | Beckman Coulter | Cat#B10247; RRID: N/A |

**Table S4.** Unique sequence counts and total template counts of TCRs identified in pre-Tx tissues. Unique sequence counts of pre-Tx detected single- or multi-tissue distributed TCRs. LLN: lung-associated lymph nodes. PBMC: peripheral blood mononuclear cells. U: unstimulated. L: CFSE^low^.

| Patient ID | Recipient pre-Tx tissues | Unique sequence counts | Total template counts | Lymphoid only unique sequence counts | Lung only unique sequence counts | PBMC only unique sequence counts | Lymphoid-lung shared unique sequence counts | Lymphoid-PBMC shared unique sequence counts | PBMC-lung shared unique sequence counts | Lymphoid-PBMC-lung shared unique sequence counts |
| --- | --- | --- | --- | --- | --- | --- | --- | --- | --- | --- |
| Pt1 | LLN_CD4_U | 67920 | 88518 | 80724 | 8825 | 68379 | 3078 | 4771 | 1839 | 3257 |
|  | LLN_CD8_U | 21333 | 104728 |  |  |  |  |  |  |  |
|  | LLN_CD4_L | 2516 | 20565 |  |  |  |  |  |  |  |
|  | LLN_CD8_L | 661 | 12287 |  |  |  |  |  |  |  |
|  | Lung | 17015 | 139407 |  |  |  |  |  |  |  |
|  | PBMC_CD4 | 72338 | 90842 |  |  |  |  |  |  |  |
|  | PBMC_CD8 | 5980 | 19652 |  |  |  |  |  |  |  |
| Pt4 | LLN_CD4_U | 214100 | 379958 | 224935 | 4062 | 55144 | 3860 | 13238 | 630 | 3120 |
|  | LLN_CD8_U | 22825 | 49985 |  |  |  |  |  |  |  |
|  | LLN_CD4_L | 7754 | 191220 |  |  |  |  |  |  |  |
|  | LLN_CD8_L | 3895 | 162298 |  |  |  |  |  |  |  |
|  | Lung | 11688 | 49298 |  |  |  |  |  |  |  |
|  | PBMC_CD4 | 49616 | 63361 |  |  |  |  |  |  |  |
|  | PBMC_CD8 | 22693 | 72016 |  |  |  |  |  |  |  |
| Pt7 | LLN_CD4_U | 290715 | 448551 | 306266 | 14279 | 36938 | 12238 | 5005 | 729 | 3141 |
|  | LLN_CD8_U | 23741 | 52735 |  |  |  |  |  |  |  |
|  | LLN_CD4_L | 13680 | 237954 |  |  |  |  |  |  |  |
|  | LLN_CD8_L | 2941 | 167535 |  |  |  |  |  |  |  |
|  | Lung | 30406 | 224480 |  |  |  |  |  |  |  |
|  | PBMC | 45851 | 61169 |  |  |  |  |  |  |  |
| Pt8 | LLN_CD4_U | 234517 | 289102 | 312474 | 8752 | 56719 | 5069 | 4365 | 304 | 1071 |
|  | LLN_CD8_U | 80256 | 191459 |  |  |  |  |  |  |  |
|  | LLN_CD4_L | 8383 | 223318 |  |  |  |  |  |  |  |
|  | LLN_CD8_L | 1079 | 33483 |  |  |  |  |  |  |  |
|  | Lung | 15242 | 34072 |  |  |  |  |  |  |  |
|  | PBMC | 62581 | 76722 |  |  |  |  |  |  |  |
| Pt9 | LLN_CD4_U | 171022 | 238144 | 222683 | 34664 | 92761 | 5443 | 24500 | 2428 | 8980 |
|  | LLN_CD8_U | 58911 | 107118 |  |  |  |  |  |  |  |
|  | LLN_CD4_L | 28464 | 350441 |  |  |  |  |  |  |  |
|  | LLN_CD8_L | 12305 | 334080 |  |  |  |  |  |  |  |
|  | Lung | 128933 | 196808 |  |  |  |  |  |  |  |
|  | PBMC | 51624 | 99372 |  |  |  |  |  |  |  |
| Pt11 | LLN_CD4_U | 275730 | 336802 | 377924 | 21285 | 210989 | 5385 | 17603 | 2187 | 5684 |
|  | LLN_CD8_U | 118868 | 169625 |  |  |  |  |  |  |  |
|  | LLN_CD4_L | 6390 | 181288 |  |  |  |  |  |  |  |
|  | LLN_CD8_L | 7570 | 400189 |  |  |  |  |  |  |  |
|  | Lung | 34585 | 75603 |  |  |  |  |  |  |  |
|  | PBMC | 236774 | 329788 |  |  |  |  |  |  |  |
| Pt12 | LLN_CD4_U | 211106 | 288940 | 231397 | 41432 | 189385 | 10367 | 18273 | 9151 | 15096 |
|  | LLN_CD8_U | 48127 | 76820 |  |  |  |  |  |  |  |
|  | LLN_CD4_L | 13913 | 335910 |  |  |  |  |  |  |  |
|  | LLN_CD8_L | 5993 | 353851 |  |  |  |  |  |  |  |
|  | Lung | 76226 | 314278 |  |  |  |  |  |  |  |
|  | PBMC | 232389 | 555188 |  |  |  |  |  |  |  |
| Pt13 | LLN_CD4_U | 150305 | 206898 | 183227 | 33630 | 117539 | 9659 | 7597 | 2428 | 6460 |
|  | LLN_CD8_U | 39383 | 82262 |  |  |  |  |  |  |  |
|  | LLN_CD4_L | 17324 | 330970 |  |  |  |  |  |  |  |
|  | LLN_CD8_L | 5613 | 279729 |  |  |  |  |  |  |  |
|  | Lung | 52273 | 99088 |  |  |  |  |  |  |  |
|  | PBMC | 134314 | 213642 |  |  |  |  |  |  |  |

**Table S5.** Absolute number and cumulative frequency of GvH and HvG CD4 and CD8 clones identified by TCRβ CDR3 DNA sequencing in pre-Tx LLN and post-Tx BAL, PBMC and TBBx in LuTx patients. “Cumulative frequency (Cum.freq.)” was calculated as a percentage of all sequences weighted by copy numbers in designated populations. LLN: lung-associated lymph nodes. BAL: bronchoalveolar lavage. TBBx: transbronchial biopsies. U: unstimulated. L: CFSE^low^. GvH: graft-versus-host. HvG: host-versus-graft.

|  |  |  | Mappable clones |  |  |  | Mappable alloreactive clones |  |  |  | Cumulative frequency of alloreactive clones |  |  |  | Total |  |
| --- | --- | --- | --- | --- | --- | --- | --- | --- | --- | --- | --- | --- | --- | --- | --- | --- |
| Patient # | T cell source | POD | D CD4 | D CD8 | R CD4 | R CD8 | GvH CD4 | GvH CD8 | HvG CD4 | HvG CD8 | GvH CD4 | GvH CD8 | HvG CD4 | HvG CD8 | Cum. freq. of GvH | Cum. freq. of HvG |
| Pt1 | LLN | 0 (L+U) |  |  |  |  | 3813 | 1692 | 2473 | 569 |  |  |  |  |  |  |
|  | LLN | 0 (U) | 87273 | 66061 | 67920 | 21333 | 491 | 213 | 268 | 194 | 0.7474 | 1.0227 | 0.7569 | 3.4079 |  |  |
|  | BAL | 64 | 200 | 271 | 306 | 198 | 22 | 7 | 16 | 6 | 0.7486 | 0.3622 | 0.6762 | 0.1449 | 1.1108 | 0.8211 |
|  | BAL | 107 | 49 | 87 | 98 | 44 | 5 | 6 | 4 | 1 | 0.8037 | 1.1481 | 0.4592 | 0.1148 | 1.9518 | 0.5741 |
|  | TBBx | 64 | 89 | 234 | 190 | 199 | 5 | 7 | 13 | 11 | 0.2218 | 0.7542 | 0.6211 | 0.8873 | 0.9760 | 1.5084 |
|  | TBBx | 107 | 84 | 199 | 147 | 156 | 8 | 8 | 0 | 8 | 0.5291 | 1.1243 | 0 | 0.5952 | 1.6534 | 0.5952 |
|  | TBBx | 177 | 139 | 214 | 202 | 202 | 13 | 5 | 11 | 14 | 0.7788 | 0.5711 | 0.6231 | 1.0384 | 1.3499 | 1.6615 |
| Pt4 | LLN | 0 (L+U) |  |  |  |  | 4895 | 2286 | 4156 | 1909 |  |  |  |  |  |  |
|  | LLN | 0 (U) | 332697 | 25963 | 214100 | 22825 | 856 | 347 | 1227 | 345 | 0.7828 | 2.3224 | 1.1414 | 2.5288 |  |  |
|  | PBMC | 320 | 50 | 10 | 6221 | 2551 | 0 | 0 | 109 | 128 | 0 | 0 | 0.4919 | 0.5530 | 0 | 1.0449 |
|  | BAL | 37 | 86 | 11 | 8781 | 3051 | 0 | 0 | 208 | 130 | 0 | 0 | 10.2361 | 16.5146 | 0 | 26.7508 |
|  | BAL | 86 | 5 | 4 | 1693 | 638 | 0 | 1 | 98 | 86 | 0 | 0.0113 | 4.1070 | 4.4116 | 0.0113 | 8.5186 |
|  | BAL | 352 | 5 | 4 | 1469 | 451 | 0 | 1 | 66 | 61 | 0 | 0.0047 | 3.3055 | 1.0623 | 0.0047 | 4.3678 |
|  | TBBx | 37 | 4 | 1 | 1589 | 487 | 0 | 0 | 35 | 25 | 0 | 0 | 4.1667 | 5.8532 | 0 | 10.0198 |
|  | TBBx | 86 | 40 | 9 | 6701 | 2311 | 0 | 0 | 18 | 9 | 0 | 0 | 1.7857 | 1.8849 | 0 | 3.6706 |
|  | TBBx | 184 | 0 | 5 | 330 | 135 | 0 | 0 | 22 | 39 | 0 | 0 | 2.9795 | 5.4004 | 0 | 8.3799 |
|  | TBBx | 268 | 1 | 0 | 170 | 73 | 0 | 0 | 19 | 19 | 0 | 0 | 1.7059 | 2.1121 | 0 | 3.8180 |
| Pt7 | LLN | 0 (L+U) |  |  |  |  | 2759 | 416 | 6298 | 1661 |  |  |  |  |  |  |
|  | LLN | 0 (U) | 186276 | 14448 | 290715 | 23741 | 523 | 146 | 1262 | 231 | 0.2808 | 1.0105 | 0.4341 | 0.9730 |  |  |
|  | PBMC | 15 | 77 | 16 | 6717 | 985 | 1 | 1 | 227 | 51 | 0.0018 | 0.0018 | 0.5659 | 0.3539 | 0.0036 | 0.9199 |
|  | PBMC | 22 | 54 | 7 | 7835 | 1000 | 1 | 0 | 246 | 64 | 0.0012 | 0 | 0.5452 | 0.2522 | 0.0012 | 0.7974 |
|  | PBMC | 56 | 64 | 8 | 8919 | 1278 | 2 | 3 | 271 | 79 | 0.0019 | 0.0253 | 0.4131 | 0.2904 | 0.0272 | 0.7034 |
|  | PBMC | 161 | 95 | 11 | 11433 | 1293 | 1 | 0 | 272 | 63 | 0.0006 | 0 | 0.2929 | 0.1992 | 0.0006 | 0.4922 |
|  | PBMC | 261 | 46 | 7 | 6942 | 1132 | 0 | 0 | 200 | 73 | 0 | 0 | 0.3814 | 0.4640 | 0 | 0.8454 |
|  | BAL | 15 | 153 | 109 | 515 | 110 | 19 | 12 | 69 | 12 | 0.5618 | 1.1827 | 3.2229 | 0.6209 | 1.7445 | 3.8439 |
|  | BAL | 22 | 77 | 85 | 737 | 228 | 3 | 7 | 135 | 27 | 0.0304 | 0.1139 | 2.9546 | 0.5772 | 0.1443 | 3.5318 |
|  | BAL | 51 | 25 | 114 | 225 | 141 | 1 | 10 | 43 | 17 | 0.0175 | 0.6132 | 1.0161 | 0.5606 | 0.6307 | 1.5767 |
|  | BAL | 176 | 27 | 133 | 1066 | 422 | 1 | 9 | 137 | 35 | 0.0066 | 0.2171 | 5.5318 | 1.1774 | 0.2236 | 6.7092 |
|  | BAL | 274 | 26 | 132 | 492 | 203 | 1 | 9 | 58 | 20 | 0.0219 | 0.8321 | 2.2115 | 0.7445 | 0.8540 | 2.9560 |
|  | TBBx | 23 | 74 | 63 | 446 | 128 | 6 | 6 | 54 | 6 | 0.1691 | 0.1972 | 1.9724 | 0.1972 | 0.3663 | 2.1696 |
|  | TBBx | 51 | 43 | 38 | 416 | 153 | 3 | 4 | 57 | 13 | 0.1427 | 0.3330 | 3.1399 | 0.6185 | 0.4757 | 3.7583 |
| Pt8 | LLN | 0 (L+U) |  |  |  |  | 6120 | 2835 | 4409 | 1008 |  |  |  |  |  |  |
|  | LLN | 0 (U) | 189767 | 140867 | 234517 | 80256 | 747 | 349 | 499 | 143 | 0.3936 | 0.2478 | 0.2128 | 0.1782 |  |  |
|  | PBMC | 6 | 72 | 88 | 3024 | 951 | 2 | 3 | 102 | 21 | 0.0034 | 0.0101 | 0.1653 | 0.6298 | 0.0135 | 0.7951 |
|  | PBMC | 15 | 149 | 88 | 6125 | 1622 | 9 | 5 | 162 | 22 | 0.0064 | 0.0042 | 0.2413 | 0.3815 | 0.0106 | 0.6228 |
|  | PBMC | 62 | 460 | 280 | 16463 | 4562 | 13 | 9 | 393 | 54 | 0.0027 | 0.0020 | 0.2250 | 0.4390 | 0.0047 | 0.6640 |
|  | PBMC | 167 | 366 | 254 | 13573 | 3167 | 13 | 6 | 236 | 36 | 0.0038 | 0.0013 | 0.1074 | 0.3228 | 0.0052 | 0.4302 |
|  | PBMC | 235 | 147 | 107 | 6988 | 1893 | 4 | 3 | 139 | 23 | 0.0035 | 0.0021 | 0.1584 | 0.3910 | 0.0057 | 0.5494 |
|  | PBMC | 356 | 75 | 52 | 5473 | 1461 | 1 | 3 | 99 | 25 | 0.0011 | 0.0033 | 0.1849 | 0.3358 | 0.0044 | 0.5207 |
|  | BAL | 7 | 135 | 120 | 446 | 236 | 17 | 4 | 46 | 10 | 0.5533 | 0.1456 | 1.6308 | 3.4362 | 0.6989 | 5.0670 |
|  | BAL | 15 | 78 | 137 | 646 | 343 | 9 | 11 | 232 | 15 | 0.1225 | 0.3184 | 6.2217 | 5.4991 | 0.4409 | 11.7208 |
|  | BAL | 56 | 65 | 153 | 415 | 238 | 3 | 10 | 67 | 9 | 0.0618 | 0.2885 | 2.1430 | 2.0400 | 0.3503 | 4.1830 |
|  | BAL | 91 | 75 | 172 | 581 | 262 | 3 | 6 | 56 | 11 | 0.0583 | 0.2138 | 1.6910 | 1.3022 | 0.2721 | 2.9932 |
|  | BAL | 154 | 66 | 168 | 614 | 256 | 5 | 5 | 35 | 13 | 0.1153 | 0.0865 | 1.3118 | 0.9226 | 0.2018 | 2.2344 |
|  | BAL | 259 | 27 | 59 | 319 | 126 | 0 | 1 | 13 | 5 | 0.0000 | 0.0494 | 1.4829 | 0.3955 | 0.0494 | 1.8784 |
|  | BAL | 294 | 63 | 120 | 465 | 159 | 2 | 0 | 23 | 8 | 0.6695 | 0.0000 | 1.3694 | 0.4869 | 0.6695 | 1.8564 |
|  | BAL | 364 | 50 | 178 | 860 | 247 | 1 | 4 | 26 | 7 | 0.0141 | 0.1124 | 1.1945 | 0.3373 | 0.1265 | 1.5318 |
|  | TBBx | 91 | 84 | 47 | 91 | 74 | 3 | 2 | 6 | 5 | 0.4994 | 0.2497 | 0.9988 | 1.1236 | 0.7491 | 2.1223 |
|  | TBBx | 294 | 78 | 47 | 73 | 36 | 3 | 0 | 3 | 1 | 0.9107 | 0.0000 | 0.5464 | 0.3643 | 0.9107 | 0.9107 |
| Pt9 | LLN | 0 (L+U) |  |  |  |  | 9201 | 7222 | 8415 | 5170 |  |  |  |  |  |  |
|  | LLN | 0 (U) | 196528 | 122773 | 171022 | 58911 | 1619 | 729 | 1329 | 523 | 1.6800 | 2.5951 | 1.3979 | 1.9194 |  |  |
|  | PBMC | 5 | 14 | 11 | 599 | 102 | 2 | 2 | 41 | 8 | 0.0784 | 0.0523 | 1.2807 | 0.2352 | 0.1307 | 1.5159 |
|  | PBMC | 21 | 67 | 48 | 6687 | 1069 | 5 | 4 | 341 | 54 | 0.0079 | 0.0095 | 1.4133 | 0.1577 | 0.0174 | 1.5710 |
|  | PBMC | 68 | 2 | 10 | 1879 | 349 | 0 | 0 | 80 | 15 | 0 | 0 | 0.9768 | 0.1431 | 0 | 1.1199 |
|  | PBMC | 158 | 267 | 180 | 18486 | 3461 | 12 | 22 | 751 | 206 | 0.0067 | 0.0096 | 1.3029 | 0.1900 | 0.0163 | 1.4928 |
|  | PBMC | 277 | 18 | 12 | 3270 | 661 | 0 | 1 | 170 | 33 | 0 | 0.0055 | 1.6751 | 0.2425 | 0.0055 | 1.9176 |
|  | PBMC | 342 | 48 | 40 | 8126 | 1541 | 0 | 3 | 374 | 84 | 0 | 0.0047 | 1.5835 | 0.2396 | 0.0047 | 1.8231 |
|  | BAL | 7 | 516 | 267 | 187 | 28 | 100 | 34 | 46 | 6 | 7.1429 | 3.6680 | 2.6544 | 0.4826 | 10.8108 | 3.1371 |
|  | BAL | 21 | 1990 | 741 | 1032 | 137 | 296 | 89 | 260 | 31 | 6.7296 | 3.0816 | 7.0091 | 0.3852 | 9.8112 | 7.3943 |
|  | BAL | 68 | 285 | 472 | 538 | 254 | 55 | 75 | 115 | 40 | 0.6051 | 2.3869 | 1.1248 | 0.3007 | 2.9919 | 1.4254 |
|  | BAL | 173 | 277 | 498 | 3024 | 461 | 44 | 72 | 203 | 31 | 1.0671 | 3.6108 | 2.0073 | 0.4153 | 4.6779 | 2.4226 |
|  | BAL | 278 | 134 | 340 | 1689 | 312 | 31 | 46 | 118 | 34 | 0.9971 | 1.9326 | 1.9120 | 1.2747 | 2.9297 | 3.1867 |
|  | BAL | 367 | 187 | 437 | 2971 | 610 | 38 | 54 | 197 | 48 | 1.8800 | 1.2054 | 1.9439 | 0.6506 | 3.0855 | 2.5945 |
|  | TBBx | 68 | 56 | 45 | 226 | 80 | 6 | 6 | 36 | 9 | 0.4205 | 0.5606 | 2.8732 | 0.7008 | 0.9811 | 3.5739 |
|  | TBBx | 89 | 10 | 9 | 175 | 69 | 0 | 0 | 18 | 4 | 0 | 0 | 2.4862 | 0.5525 | 0.0000 | 3.0387 |
|  | TBBx | 173 | 2 | 1 | 195 | 41 | 1 | 0 | 15 | 3 | 0.1969 | 0 | 3.1496 | 0.5906 | 0.1969 | 3.7402 |
|  | TBBx | 202 | 86 | 78 | 1116 | 197 | 8 | 12 | 86 | 17 | 0.3161 | 0.4109 | 3.1922 | 0.7269 | 0.7269 | 3.9191 |
|  | TBBx | 278 | 7 | 31 | 976 | 292 | 1 | 4 | 64 | 24 | 0.0289 | 0.2888 | 2.4256 | 1.1551 | 0.3176 | 3.5807 |
|  | liver Bx | 5 | 316 | 181 | 931 | 158 | 54 | 14 | 138 | 27 | 1.8260 | 0.7101 | 4.8948 | 1.7246 | 2.5361 | 6.6193 |
|  | liver Bx | 81 | 3 | 5 | 468 | 347 | 0 | 1 | 67 | 33 | 0 | 0.0245 | 2.4205 | 1.1002 | 0.0245 | 3.5208 |
| Pt11 | LLN | 0 (L+U) |  |  |  |  | 2883 | 1815 | 3817 | 4280 |  |  |  |  |  |  |
|  | LLN | 0 (U) | 222042 | 46584 | 275730 | 118868 | 682 | 133 | 611 | 253 | 0.5820 | 1.6785 | 0.4792 | 0.9155 |  |  |
|  | PBMC | 8 | 441 | 175 | 16284 | 4025 | 6 | 8 | 359 | 160 | 0.0022 | 0.0034 | 0.2654 | 0.4217 | 0.0056 | 0.6872 |
|  | PBMC | 36 | 372 | 177 | 14775 | 4030 | 6 | 9 | 305 | 152 | 0.0027 | 0.0150 | 0.1852 | 0.8093 | 0.0177 | 0.9945 |
|  | PBMC | 71 | 753 | 276 | 28149 | 6041 | 12 | 13 | 468 | 224 | 0.0016 | 0.0050 | 0.2018 | 0.4039 | 0.0066 | 0.6057 |
|  | PBMC | 167 | 320 | 127 | 16685 | 3910 | 7 | 9 | 324 | 143 | 0.0019 | 0.0118 | 0.2219 | 1.0237 | 0.0137 | 1.2456 |
|  | PBMC | 440 | 344 | 123 | 18004 | 4956 | 4 | 10 | 346 | 163 | 0.0010 | 0.0021 | 0.2525 | 0.7865 | 0.0030 | 1.0390 |
|  | BAL | 28 | 294 | 269 | 140 | 112 | 16 | 23 | 26 | 11 | 0.6587 | 2.7917 | 1.5997 | 0.6587 | 3.4504 | 2.2585 |
|  | BAL | 163 | 552 | 454 | 1443 | 673 | 16 | 22 | 74 | 44 | 0.1154 | 0.4916 | 0.4565 | 0.6872 | 0.6069 | 1.1437 |
|  | BAL | 387 | 146 | 204 | 421 | 391 | 1 | 9 | 18 | 20 | 0.0148 | 0.3562 | 0.2968 | 1.0092 | 0.3710 | 1.3060 |
|  | TBBx | 28 | 233 | 202 | 34 | 36 | 7 | 12 | 2 | 4 | 0.6426 | 1.4458 | 0.1606 | 0.3213 | 2.0884 | 0.4819 |
|  | TBBx | 77 | 490 | 308 | 755 | 389 | 9 | 9 | 33 | 19 | 0.1286 | 0.3858 | 0.5715 | 0.5429 | 0.5144 | 1.1144 |
|  | TBBx | 128 | 2 | 18 | 6 | 8 | 0 | 0 | 0 | 1 | 0 | 0 | 0 | 1.1111 | 0 | 1.1111 |
|  | TBBx | 163 | 366 | 319 | 331 | 201 | 11 | 16 | 43 | 17 | 0.2526 | 1.0686 | 1.0297 | 0.6412 | 1.3212 | 1.6709 |
| Pt12 | LLN | 0 (L+U) |  |  |  |  | 5592 | 3472 | 5236 | 2458 |  |  |  |  |  |  |
|  | LLN | 0 (U) | 217758 | 59664 | 211106 | 48127 | 1565 | 205 | 917 | 159 | 2.0143 | 0.9212 | 0.9140 | 0.9529 |  |  |
|  | PBMC | 10 | 61 | 25 | 6704 | 1771 | 1 | 0 | 234 | 45 | 0.0018 | 0.0000 | 0.8021 | 0.1622 | 0.0018 | 0.9643 |
|  | PBMC | 49 | 184 | 93 | 18131 | 3706 | 4 | 4 | 454 | 93 | 0.0024 | 0.0016 | 0.6852 | 0.1811 | 0.0040 | 0.8662 |
|  | PBMC | 112 | 238 | 91 | 20592 | 4338 | 2 | 6 | 487 | 107 | 0.0006 | 0.0020 | 0.6634 | 0.2128 | 0.0025 | 0.8762 |
|  | PBMC | 196 | 127 | 41 | 12307 | 2898 | 0 | 1 | 312 | 66 | 0 | 0.0007 | 0.6845 | 0.1981 | 0.0007 | 0.8826 |
|  | PBMC | 315 | 64 | 26 | 8288 | 2131 | 1 | 1 | 207 | 46 | 0.0011 | 0.0011 | 0.5511 | 0.1583 | 0.0023 | 0.7094 |
|  | BAL | 10 | 386 | 181 | 1571 | 597 | 79 | 14 | 258 | 35 | 1.4544 | 0.3458 | 12.5305 | 1.0069 | 1.8002 | 13.5374 |
|  | BAL | 106 | 65 | 82 | 140 | 93 | 6 | 2 | 20 | 4 | 0.6179 | 0.2060 | 2.5747 | 0.4119 | 0.8239 | 2.9866 |
|  | BAL | 309 | 94 | 88 | 428 | 243 | 11 | 5 | 37 | 12 | 0.5367 | 0.2013 | 2.5159 | 1.2076 | 0.7380 | 3.7236 |
|  | TBBx | 55 | 63 | 25 | 87 | 75 | 10 | 1 | 4 | 1 | 1.9064 | 0.1733 | 1.0399 | 0.1733 | 2.0797 | 1.2132 |
|  | TBBx | 106 | 1088 | 304 | 526 | 318 | 152 | 8 | 67 | 34 | 5.2106 | 0.2402 | 1.9217 | 0.9424 | 5.4508 | 2.8640 |
| Pt13 | LLN | 0 (L+U) |  |  |  |  | 7957 | 8406 | 6115 | 2398 |  |  |  |  |  |  |
|  | LLN | 0 (U) | 201323 | 56882 | 150305 | 39383 | 1034 | 181 | 1231 | 661 | 1.1068 | 0.5740 | 1.6955 | 6.6945 |  |  |
|  | PBMC | 7 | 399 | 184 | 15322 | 6535 | 18 | 28 | 670 | 670 | 0.0048 | 0.0066 | 0.3273 | 1.9549 | 0.0114 | 2.2822 |
|  | PBMC | 23 | 431 | 179 | 15999 | 5988 | 18 | 31 | 755 | 596 | 0.0048 | 0.0084 | 0.3445 | 1.2695 | 0.0132 | 1.6141 |
|  | PBMC | 98 | 279 | 127 | 13041 | 5409 | 5 | 20 | 575 | 573 | 0.0015 | 0.0066 | 0.3299 | 1.3404 | 0.0080 | 1.6703 |
|  | PBMC | 190 | 10 | 4 | 805 | 793 | 0 | 0 | 38 | 82 | 0 | 0 | 0.4878 | 1.8069 | 0 | 2.2946 |
|  | BAL | 8 | 628 | 205 | 1310 | 885 | 195 | 30 | 379 | 128 | 2.4486 | 0.4948 | 6.8680 | 6.3902 | 2.9434 | 13.2583 |
|  | BAL | 21 | 489 | 234 | 2758 | 2941 | 156 | 28 | 624 | 384 | 0.2843 | 0.1055 | 4.8757 | 24.0390 | 0.3898 | 28.9146 |
|  | BAL | 50 | 49 | 77 | 439 | 578 | 8 | 6 | 104 | 65 | 0.2280 | 0.4053 | 4.5593 | 4.7366 | 0.6332 | 9.2958 |
|  | BAL | 106 | 50 | 141 | 337 | 392 | 11 | 19 | 82 | 52 | 0.4083 | 1.4447 | 4.6796 | 2.8266 | 1.8530 | 7.5063 |
|  | BAL | 197 | 329 | 326 | 1517 | 1566 | 104 | 32 | 189 | 188 | 0.2686 | 0.3035 | 0.7146 | 1.3504 | 0.5720 | 2.0650 |
|  | TBBx | 50 | 38 | 25 | 595 | 779 | 6 | 0 | 83 | 87 | 0.1214 | 0 | 2.2060 | 3.1370 | 0.1214 | 5.3430 |

**Table S6.** Summary of the unique sequence numbers of each type of microbial-reactive TCRs compiled from published databases.

| Abbreviation | Full name | Number of unique TCRβ sequences compiled |
| --- | --- | --- |
| CMV | Cytomegalovirus | 209,511 |
| EA | Enterobacter aerogenes | 159 |
| EBV | Epstein-Barr virus | 4,979 |
| E.Coli | Escherichia Coli | 394 |
| Flu | Influenza | 8,215 |
| HCV | Hepatitis C virus | 662 |
| HIV | Human immunodeficiency virus | 3,339 |
| KP | Klebsiella Pneumoniae | 323 |
| MTB | Mycobacterium tuberculosis | 30,958 |
| SA | Staphylococcus aureus | 1,877 |
| ST | Salmonella typhimurium | 26,477 |
| Shi | Shigella | 318 |
| SarsCoV-2 | Severe acute respiratory syndrome Coronavirus 2 | 148, 679 |
| YFV | Yellow fever virus | 746 |

## Notes

### Author Declarations

Ethics committee/IRB of the Columbia University gave ethical approval for this work.

### Summary of Updates

Introduction on lung transplantation was updated with 2025 data. Table S1 was updated with recent death events. A co-author (V. M.) missed from the last submission was added back.

